# On the Proportion of Patients Who Experience a Prodrome Prior to Psychosis Onset - A Systematic Review and Meta-analysis

**DOI:** 10.1101/2023.05.15.23290015

**Authors:** David Benrimoh, Viktor Dlugunovych, Abigail C Wright, Peter Phalen, Melissa C. Funaro, Maria Ferrara, Albert Powers, Scott Woods, Sinan Guloksuz, Alison R Yung, Vinod Srihari, Jai Shah

## Abstract

**BACKGROUND:** Preventing or delaying the onset of psychosis requires identification of those at risk for developing psychosis. For predictive purposes, the prodrome – a constellation of symptoms which may occur before the onset of psychosis – has been increasingly recognized as having utility. However, it is unclear what proportion of patients are expected to experience a prodrome or how this varies according to the definition used.

**METHODS:** We conducted a systematic review and meta-analysis of studies of patients with psychosis with the objective of determining the proportion of patients who experienced a prodrome prior to psychosis onset. Inclusion criteria included a consistent prodrome definition and reporting the proportion of patients who experienced a prodrome. We excluded studies of only patients with a prodrome or solely substance-induced psychosis, qualitative studies without prevalence data, conference abstracts, and case reports/case series. We searched Ovid MEDLINE, Embase (Ovid), APA PsycInfo (Ovid), Web of Science Core Collection (Clarivate), Cochrane Database of Systematic Reviews, Cochrane Central Register of Controlled Trials, APA PsycBooks (Ovid), ProQuest Dissertation & Thesis, on March 3, 2021. Studies were assessed for quality using the Critical Appraisal Checklist for Prevalence Studies. Narrative synthesis and proportion meta-analysis were used to estimate prodrome prevalence. I^2^ and predictive interval were used to assess heterogeneity. Subgroup analyses were used to probe sources of heterogeneity. (PROSPERO ID: CRD42021239797).

**RESULTS:** Seventy-one articles were included, representing 13,774 patients. Studies varied significantly in terms of methodology and prodrome definition used. The random effects proportion meta-analysis estimate for prodrome prevalence was 78.3% (95% CI= 72.8-83.2); heterogeneity was high (I^2^ 97.98% [95% CI= 97.71-98.22]); and the prediction interval was wide (95% PI= 0.411-0.936). There were no meaningful differences in prevalence between grouped prodrome definitions, and subgroup analyses failed to reveal a consistent source of heterogeneity.

**CONCLUSIONS:** This is the first meta-analysis on the prevalence of a prodrome prior to the onset of first episode psychosis. The majority of patients (78.3%) were found to experience a prodrome prior to psychosis onset. However, findings are highly heterogenous across study and no definitive source of heterogeneity was found. As most studies were retrospective in nature, recall bias likely affects these results. While the large majority of patients with psychosis experience a prodrome in some form, it is unclear if the remainder of patients experience no prodrome, or if ascertainment methods employed in the studies were not sensitive to their experiences. Given widespread investment in indicated prevention of psychosis through prospective identification and intervention during the prodrome, a resolution of this question as well as a consensus definition of the prodrome is much needed in order to effectively direct services, and may be accomplished through novel, densely sampled prospective cohort studies.

## INTRODUCTION

Schizophrenia spectrum psychotic disorders affect close to 1% of the population globally and are significant drivers of disability and healthcare costs (Kessler et al., 2005; Wu et al., 2006; Desai et al., 2013; Saha et al., 2005; Moreno-Küstner et al., 2018). With a view to improving prognostics and early intervention, there has long been an interest in characterizing the onset and early course of psychosis – including risk, premorbid, and sub-threshold periods (Hafner et al., 2003). Initial investigations centering on patients who had already developed psychosis were focused on examinations of the pre-psychotic period known as the *prodrome*, which involved collection of retrospective data. In a seminal study by Hafner et al. (1999), the prodrome was described as a period of symptoms (including, but not limited to, changes in affect, cognition, and social behavior) that was contiguous with the onset of psychosis: as such, a prodrome could only be retrospectively identified (once a psychosis had occurred), and was identified present in 73% of psychotic patients. Indeed, many models of illness development indicate the number of often nonspecific symptoms, negative symptoms, so-called “basic symptoms”, and mood changes preceding the onset of positive psychotic symptoms (e.g. Gross et al., 1987; McGlashan et al., 1996; Cupo et al., 2021).

Via the early intervention movement, the opportunity to prevent or delay onset of psychosis presented researchers with a need to prospectively identify individuals at risk for developing psychosis. Focusing on milder or “sub-threshold” versions of the characteristic symptoms of a full-blown disorder (IOM, 1994), prevention efforts therefore highlighted attenuated or brief intermittent psychotic symptoms such as perceptual abnormalities, subthreshold hallucinations or delusions, disorganization of speech and odd or unusual behavior (Yung et al., 2005; McGlashan et al., 2001). The resulting “clinical high-risk”(CHR; also known as the at-risk mental state [ARMS] or ultra high-risk [UHR]) state thus represents a ‘putative’ prodrome which prospectively identifies those close-in to the point of transitioning to psychosis who could then be followed longitudinally (McGorry et al., 2003).

Studies have now demonstrated that help-seeking individuals with these symptoms have an elevated risk of transition to psychosis for up to 10 years (Fusar-Poli et al., 2012; Salazar de Pablo et al., 2021a). These prospective definitions have also assisted in the development of novel service offerings - early intervention clinics, aimed at providing care to patients experiencing CHR states (Salazar de Pablo et al., 2021b). Work focusing on CHR has garnered much excitement, demonstrating evidence of effectiveness as measured by reductions in duration of untreated psychosis, improvement of symptomatic and functional outcomes, while being cost-effective (Killackey & Yung, 2007; Devoe et al., 2020), though there is more limited evidence for reduction of rates of CHR symptoms and of transition to psychosis (Fusar-Poli et al., 2019; Davies et al., 2018; Worthington & Cannon, 2021).

Despite the success of work focusing on the CHR syndrome, emerging evidence regarding trajectories to psychosis has further textured our understanding of the role of the CHR syndrome and its relationship with the prodrome. First, it is now clear that the majority of CHR cases do not go on to develop psychosis, even up to 10 years following initial identification of an at-risk state (Fusar-Poli et al., 2012; Gale et al., 2013; Addington et al., 2020). Second, despite the fact that most CHR patients do not transition to psychosis, they nonetheless have high rates of developing nonpsychotic mental disorders - suggesting that ‘heterotypic’ shifts across diagnostic categories are frequent in this population (Simon et al.,, 2001; Addington et al., 2017; though this is not always demonstrated (Lin et al., 2015; Woods et al., 2018). Third, follow-back studies have now reported that in a minority of first episode psychosis cases, no identifiable pre-onset subthreshold psychotic symptoms (representing a CHR state) could be found - even when using a broad definition of prodrome (Shah et al., 2017; Schultze-Lutter et al., 2015). Even if such cases in fact represent a rapid onset of psychosis in which the at-risk state appears only momentarily before transitioning to FEP, this reduces (for those subjects) the period during which early identification aimed at the CHR stage might be effective.

In light of increasing research and programmatic investment in the CHR phase (Brady et al., 2023), these data highlight the need to consolidate knowledge regarding the question of what proportion of patients who develop psychosis actually experience a prior prodrome and/or prior sub-threshold positive symptoms (hereinafter referred to as the prodrome). Such information would be immediately relevant for determining the upper limit of how diagnostically-bounded CHR services alone can address the population of patients who will ultimately develop psychosis, either at present or if their reach is extended (Ajnakina et al., 2019). Alternately, it could generate novel models of service design, delivery or integration to delay or prevent heterotypic trajectories to psychosis. Inconsistency in measured prodrome prevalence may also be linked to changes in how the prodrome is conceptualized and captured across different research approaches (e.g. prospective versus retrospective) and definitions (e.g. broad symptoms versus sub-threshold psychotic symptoms), and might in turn inform how such definitions can be applied in the future across clinical and research settings. Better understanding of prodrome prevalence definition and variability across studies may also help to make progress in identifying differences in prodromal phenotypes - including the absence of a prodrome - which may reflect different underlying neurobiological mechanisms, the study of which may yield useful biomarkers.

We therefore sought to fill this gap in the literature by conducting a systematic review and meta-analysis of studies to determine what proportion of patients experience a prodrome prior to psychosis onset. In keeping with the result of Hafner et al, we hypothesized that the majority of patients- in excess of 70%- would have a variably-defined prodrome prior to psychosis onset.

We also expected the definitions of prodrome to vary considerably within the literature, and for broader definitions of the prodrome (that included more symptoms) to result in higher prodrome prevalences.

## METHODS

The reporting of this systematic review and meta-analysis was guided by the Preferred Reporting Items for Systematic Review and Meta-Analysis (PRISMA) statement. (Page et al 2020) and was pre-registered on PROSPERO (see supplementary for details). There were no deviations from the published protocol other than the addition of proportion meta-analysis (Barker et al., 2021) as an analytic technique.

### Research Question

Our main research question was: “what proportion of patients who develop psychosis experience a prodromal phase prior to psychosis onset?” Because there have been different definitions of what a prodrome *is* over time, a secondary question was “how do the variable definitions and methods of measuring the prodrome affect the proportion of patients who experience a prodrome”? With respect to prodrome definitions, there is currently no gold-standard definition of a prodrome and we had no *a priori* reason to select one definition over another. We therefore adhered to the definition articulated in each study, and sought to quantify the inconsistency of results reported in the literature.

### Inclusion and Exclusion Criteria

Inclusion criteria were as follows: 1) studies of first episode or subsequent episode psychosis (both affective and non-affective were included, as well as psychosis not otherwise specified) in which the prevalence of prodromal symptoms was established (whether the *primary* aim of the study or not) or 2) studies of general population cohorts followed prospectively to determine how many people experience a prodrome and eventual psychosis. In addition, studies had to 3) be studies of populations of patients, 4) provide the proportions of people who experienced a prodrome (as defined by the study) prior to onset of psychosis, and 5) apply a consistent definition of the prodrome within the study. This definition could range from specific (e.g. meeting a threshold on a specific scale) to general (e.g. a brief description of symptoms), as long as it was consistently applied.

Exclusion criteria were as follows: 1) studies in which experiencing a prodrome was an inclusion criterion as patients who developed psychosis in these cohorts would, by definition, have had a preceding prodrome, artificially inflating the proportion to 100%. We also excluded 2) qualitative studies that did not report prevalence data as well as protocols, conference proceedings/abstracts, reviews, and case studies/case series; and 3) studies solely of patients with substance-induced psychosis (though studies with a minority of patients with drug-induced psychosis were allowed; it was generally not possible to separate these patients out in prevalence calculations). Further details regarding inclusion/exclusion criteria can be found in supplementary methods.

### Search Strategy

On March 3, 2021 a comprehensive search was conducted using electronic databases: Ovid MEDLINE, Embase (Ovid), APA PsycInfo (Ovid), Web of Science Core Collection (Clarivate), Cochrane Database of Systematic Reviews, Cochrane Central Register of Controlled Trials, APA PsycBooks (Ovid), and ProQuest Dissertation & Thesis. No date or language filters were used. Unpublished studies, or “gray literature” (e.g. theses, program evaluation) was also included. All search strategies are presented in the supplementary material.

The final search retrieved a total of 12852 references, which were pooled in EndNote 20 and deduplicated by the Reference Deduplicator (Yale, 2021). This set was uploaded to the Covidence (Covidence, 2023) platform for article selection. Four articles not identified by the search were added in on the advice of experts in the field. Three of these articles did not have appropriate keywords in the abstract and title (Varsamis et al., 1971; Huber et al., 1980; Häfner et al., 1989) and one of them was published after our search had been conducted (Ferrara et al., 2021). Given the significant amount of time required to process the articles, the decision was made not to update the search once the data extraction was complete and to defer this to an updated review in the future. A flowchart per PRISMA is presented in Figure 1.

**Figure 1:**
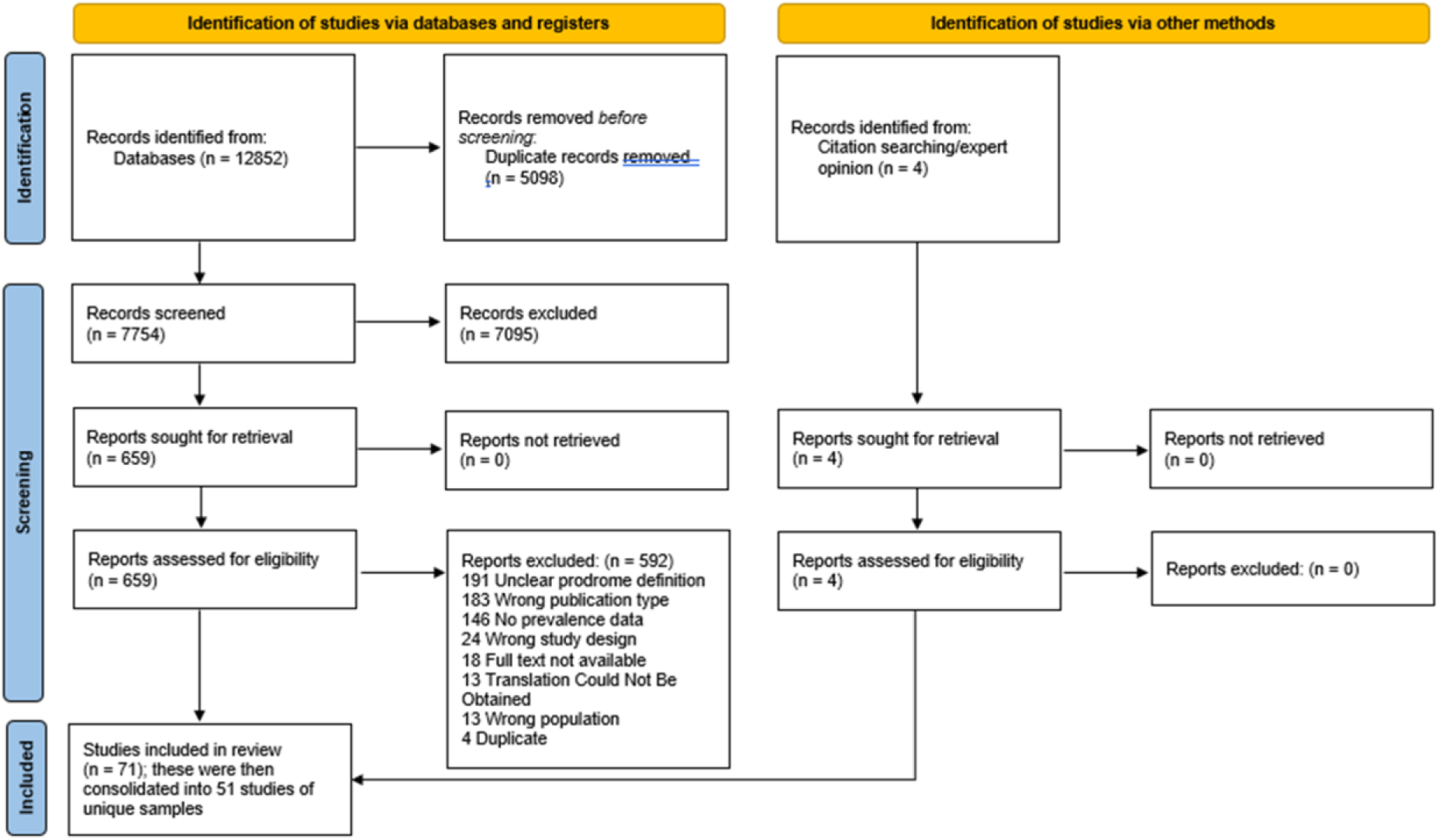
Prisma Diagram [High Quality version attached]

### Article Selection

Each article was screened by title and abstract by two independent reviewers (D.B., A.C.W., V.D., P.P.); conflicts were resolved by an expert in the field (J.S.). Included articles were then subject to a full text review by two independent raters (D.B., A.C.W., V.D., P.P.); conflicts were resolved by group consensus at meetings including J.S.

### Translations and Requests for Missing Data

For non-English articles, native speakers of the language in question with relevant expertise were sought out to assist with extraction. Speakers of English, French, Russian, Polish, and Italian were available. When these native non-English language speakers could not be found, the DEEP-L translation service (www.deepl.com) was used to provide article translations. When this translation failed or produced an unreadable article, the paper was excluded. Where further information was deemed necessary, we attempted to contact the corresponding authors of 53 articles for data or clarifications of prodrome definition and received 19 responses.

### Data Extraction

Data extraction of study and patient characteristics as well as prodrome prevalence proceeded using a standardized form (available in supplementary methods). In studies which included both participants with and without psychosis, prevalence was assessed based on the total sample with psychosis. Data was extracted by one primary reviewer and this extraction was validated by a second reviewer. Conflicts in extraction were resolved via group discussion involving (D.B., V.D., A.C.W., and J.S.).

Once extracted, data was consolidated into a final data table, a subset of which is presented as Table 1. If several articles reported on the same sample, they were presented as a single entry in Table 1 and considered as a single datapoint reflected in the PRISMA diagram (Fig. 1).

**Table 1:** Included Studies [provided in attachment]

### Assessment of Article Quality

Quality was assessed using the Critical Appraisal Checklist for Prevalence Studies published by the Joanna Briggs Institute (hereinafter referred to as the JBI) (Martin 2017). Because of our interest in heterogeneity, we did not exclude articles deemed to be of poor quality: instead of the JBI checklist item asking the reviewer to decide to include or exclude the article, we modified the scale by asking each reviewer to independently rate the article as being of “good”, “fair” or “poor” quality based on their overall assessment of the checklist criteria. To conservatively estimate study quality, the lower of the two reviewers’ ratings was assigned to the article.

### Grouping for analyses

The primary analysis of prodrome prevalence and literature heterogeneity (I^2^ and prediction interval; Higgins et al., 2003; Migliavaca et al., 2022) included all selected and extracted studies. We also report the prediction interval (the interval in which the prevalence estimate from the next hypothetical study to be added to the metanalysis is expected to lie) for the prevalence of prodrome to supplement our estimate of heterogeneity (Spence et al., 2016; Migliavaca et al., 2022). To determine potential sources of heterogeneity in the primary analyses and to assess the impact of measurement approaches, study population selection, or methodology on prodrome prevalence, further estimates of I^2^, prediction interval and prevalence were performed on the following subgroups as secondary and exploratory analyses: only those studies rated as being “fair” or “good” on quality assessment; only those studies conducted within first episode psychosis clinics; only those studies conducted using patient interviews; only those studies deriving estimates from self report; only those studies conducted on a population or catchment area sample; only those studies conducted using chart review; studies which included solely inpatients; studies with definitions inspired by the Interview for the Retrospective Assessment of the Onset of Schizophrenia (IRAOS; a detailed symptom-based measure) (Hafner et al., 1992); studies with definitions inspired by the DSM-III (APA, 1980) (which is syndromal in nature as opposed to being focused on individual symptoms); studies including only patients with schizophrenia spectrum disorders; studies including patients with more heterogeneous diagnoses (e.g. including affective psychosis and delusional disorder and other psychotic disorders); and only those studies which used a validated prodrome scale. Subgroup analyses are recommended in proportion meta-analysis, as they can assist in the determination of sources of heterogeneity (Migliavaca et al., 2022). In cases where a study’s membership in a subgroup was unclear, it was assumed not to be part of the subgroup.

### Meta-analysis

For our primary aim of determining the prevalence of prodrome prior to psychosis onset, a meta-analysis of reported proportions was conducted. We used random effects models given the expected inconsistency between studies in terms of results and methodology and present the results as a forest plot. Meta-analyses were carried out using MedCalc v20.2 (MedCalc Software Ltd.). Heterogeneity (the variation in estimates between studies, whether in primary or subgroup analyses) was assessed using the I^2^ metric (Higgins et al., 2003) as well as the prediction interval. The prediction interval, which assesses the interval within which a new point estimate would lie based on the studies in the meta-analysis, and which provides another estimate of data variability with clinical relevance (based knowledge of what would constitute clinically relevant uncertainty), was calculated using Comprehensive Meta-Analysis Version 4 (Migliavaca et al., 2022; Borenstein et. al., 2022).

### Publication bias

The presence of publication bias was assessed using the Begg’s test and funnel plot (Begg and Mazumdar, 1994). Given the relative lack of commercial interests in this field, we did not expect there to be significant publication bias.

### Categorizing prodrome definitions

We grouped prodrome definitions into three categories as follows. The first was the “Non-specific” group, which consisted of those studies which had brief or underspecified definitions; for example one study in this category defined the prodrome as a “disturbance or deviation from the patient’s previous experience and behavior that occurs before the development of florid psychotic features” (Tan et al., 2001); these may have, but did not always, include attenuated psychotic symptoms. A further example of the “Non-specific” group would be the study by Dominguez-Martinez et al., 2017, who defined the onset of the prodrome as “the earliest clinically significant deviation from the patient’s premorbid personality… established considering the first appearance of either attenuated positive or negative symptoms”. This was judged as being non-specific because any number of symptoms could be considered as fulfilling these criteria.

The second group was the “Attenuated Psychotic Symptoms Only” or “APS Only” group; this group defined the prodrome solely on the basis of sub-threshold psychotic symptoms such as perceptual changes or the onset of bizarre thoughts (e.g. Shah et al., 2017, where the focus was on 9 expert-defined sub-threshold psychotic symptoms, and Ferrara et al., 2021 where the symptoms were defined based on the Structured Interview for Psychosis-Risk Syndromes (SIPS) assessment (Miller et al., 2002)).

The third group was the “Specified Broad” group. This group considered explicit lists of symptoms or diagnoses (as opposed to the “Non-specific” group) which were broader than (but could nonetheless include) APS. An example of the “Specified Broad” category would be the Hafner et al., 1995 study, in which a specific instrument (the IRAOS) was used to establish the presence of a number of specified symptoms. These groups are presented in Table 2.

**Table 2:** Prodrome definitions: studies and sample sizes [provided in attachment]

Further exploratory analyses were conducted to assess the impact of changing definitions over time and regions on prevalence rates and are presented in the supplementary material.

## RESULTS

### Articles selected

Results of the article selection process are demonstrated in the PRISMA flow diagram (Fig. 1). The search resulted in 12,852 articles. After removal of duplicates, 7,758 studies were screened. 663 relevant studies were assessed as full texts, of which 592 were excluded, leaving 71 articles in the review. The three most common reasons for exclusion were: unclear (or missing) definition of the prodrome which did not allow us to assess what the authors meant by the prodrome; ineligible type of publication (e.g. a conference abstract); or article did not contain prodrome prevalence data. Note that a given article may have had multiple reasons for exclusion, but only one reason for exclusion, based on a structured and ordered list agreed by the extraction team, was recorded per article. After merging articles which dealt with the same samples, our final dataset for this review included 51 studies.

Included studies are described in Table 1. Twenty-one studies (41.2%) were conducted in Europe; 15 (29.4%) in North America; 9 (17.6%) in Asia; 4 (7.8%) in Oceania; and 2 (3.9%) were conducted in multiple countries. These regions are of course not homogeneous with respect to language, ethnicity, culture, medical practices, and a host of other variables, but are grouped to facilitate analysis. There were no studies from South America or Africa. The majority of studies were conducted at specialty clinics, university-affiliated sites, hospitals, or within urban areas, indicating a lack of representation from community and rural sites; this is counterbalanced by other large studies examining large population samples and primary care/community clinics.

For the 44 studies that reported detailed sex or gender data, the average percentage of a sample that was male was 64% (SD=0.13). All but 4 studies were published after 1980, the year the DSM-3 was released (APA, 1980).

With respect to quality, 30 (58.8%) studies were of “Fair” quality, 7 (13.7%) of “Good” quality, and 14 (27.5%) of “Poor” quality. Methodologically, 19 (37.3%) of studies did not use a validated prodrome scale; 41 (80.4%) of studies included some form of interview with the patient; 6 (11.8%) relied solely on chart review (one of these used administrative diagnosis data rather than the chart itself); and 3 (5.9%) relied solely on self-report (i.e. questionnaires completed by patients). Forty nine (96.1%) studies determined the presence of the prodrome in a retrospective fashion (i.e. follow-back analyses after psychosis onset, relying on patient and family recall or on documentation available from before onset). There was one clearly prospective study, Maki et al, 2014, where patients from a birth cohort were administered a prodrome screening questionnaire during one time interval and were then followed to determine whether they developed psychotic symptoms during a later time interval.

Eighteen (35.3%) of the studies were catchment-based or population-level studies. With respect to setting, 24 (47%) of studies recruited solely inpatients and 9 (17.6%) were conducted at first episode psychosis/first episode schizophrenia clinics. The most common diagnoses (Table 1) were schizophrenia and schizophrenia spectrum disorders.

### Prodrome prevalence

The combined sample size of the included studies was 13,774 patients with psychosis. The primary outcome of this review is an estimate of the prevalence of the prodrome prior to psychosis onset. Note that for one study (Jackson et al., 1995) the authors offered several prevalences according to varying definitions; we selected the definition that produced the highest prevalence for the purposes of the meta-analysis. The results are demonstrated in the forest plot Fig 2a and the funnel plot in Fig 2b (full weights per study are available in the supplementary results).

**Figure 2a:**
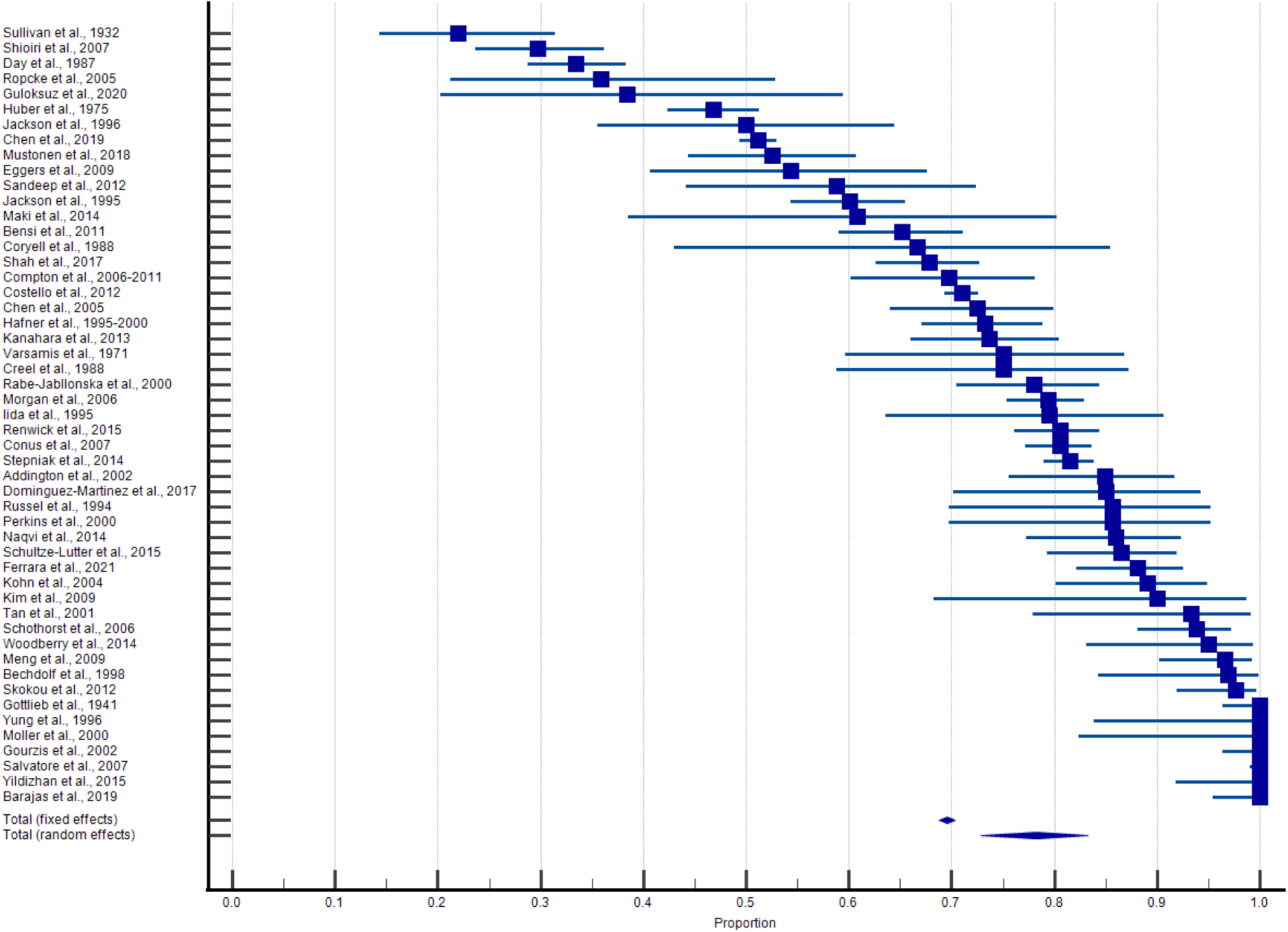
Forest Plot for all Studies [High Quality version attached]

**Figure 2b:**
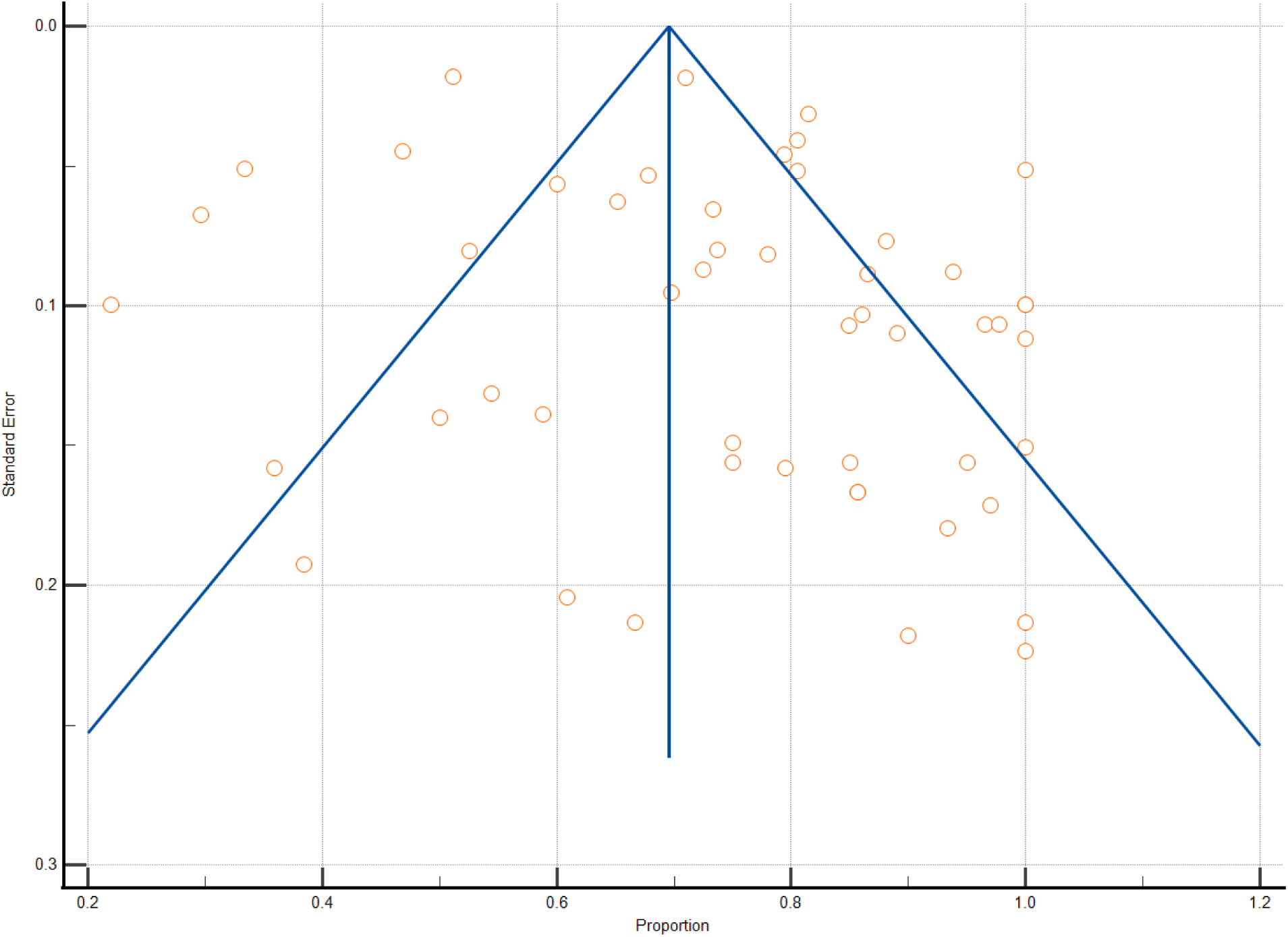
Funnel Plot for All Studies [High Quality version attached]

The random effects meta-analysis estimate of the prodrome prevalence is 78.3% (95% CI= 72.8-83.2): included studies found that 78.3% of patients with psychosis experienced a pre-

onset prodrome of one definition or another. The I^2^ for this analysis is 97.98% (95% CI= 97.71- 98.22), demonstrating high inconsistency. The prediction interval was wide (95% PI= 0.411- 0.936). Consistent with the funnel plot, there was a low risk of publication bias on Begg’s test (Kendall’s Tau= 0.015; *p* = 0.88).

### Prodrome definitions

There was relatively little variation in prodrome prevalence across definition categories, contrary to our initial hypothesis. Seventeen (33.3%) of the studies fell into the “Non-specific” definition category; these studies had a mean prevalence of 76.9% (SD=20.4%). Four studies (7.8%) fell into the “APS only” definition category; these studies had a mean prevalence of 72.3% (SD= 25.4%). Thirty (58.8%) studies fell into the “Specified Broad” category; these had a mean prevalence of 74.5% (SD=21.2%). We included in this latter category the study by Costello (2012) which focused on administrative data, since the authors specified that any preceding mental health diagnoses would be considered to be prodromal. Final groupings based on definition can be found in table 2.

### Prodrome prevalence - subgroup analyses

Given the high inconsistency present in the estimate of prodrome prevalence derived from all the studies, we conducted multiple post-hoc subgroup analyses aimed at identifying potential sources of heterogeneity. These results are presented in Table 3. No subgroups demonstrated publication bias (Begg’s test *p*’s all > 0.05).

**Table 3:** Meta-analysis Results for Subgroups [provided in attachment]

As can be seen when reviewing Table 3, the majority of the subgroups had point estimates close to the overall estimate of 78.3%. Nonetheless, the individual estimates from each subgroup have high heterogeneity and wide prediction intervals. More intensive data gathering methodologies (e.g. interviews, using a prodrome scale) and the use of validated instruments did tend to generate higher prodrome prevalence than studies with less intensive methodologies (e.g. chart review, self-report). Overall, studies generated similar estimates even when using different definitions and when using instruments with different approaches to determining if the prodrome had been present (e.g. the DSM-III vs. the IRAOS).

Furthermore, with the exception of self report, the degree of inconsistency within each subgroup remained extremely high in all subgroups. We note that the estimates for the two studies using the SIPS assessment were relatively close: 88.1% in Ferrara et al., 2021 and 95% in Woodberry et al., 2014, but the inconsistency within the APS group is high when including all APS-definition studies. The FEP service-only subgroup yielded an estimate close to the group average; as such, the shorter recall times theoretically afforded by focusing on FEP patients does not appreciably affect the estimate. Finally, the small self-report subgroup has both a much lower estimate of the prevalence (54.8%) as well as a much lower inconsistency (0%) compared to both the analysis including all data as well as every other subgroup; however, this inconsistency had a wide confidence interval, indicating that the I^2^ estimate for this subgroup (which contains only four studies) is uncertain. Predictive intervals in the subgroups were also generally wide.

## DISCUSSION

This is the first systematic review and meta-analysis to determine what proportion of patients experience a prodromal phase prior to onset of threshold-level psychosis. Our results confirm the results of previous work (Table 1) that a prodrome is experienced by a substantial majority of patients who develop psychosis. Our overall estimate is a prevalence of 78.3%, though individual studies have reported prevalences as low as 22% and as high as 100% - representing high heterogeneity (I^2^ 97.98% [95% CI= 97.71-98.22]) and a wide prediction interval (95% PI= 0.411-0.936). There were no meaningful differences in prevalence between grouped prodrome definitions, and subgroup analyses failed to reveal a consistent source of heterogeneity. Implicit in the question of how frequently the prodrome occurs before psychosis, however, are two assumptions that deserve to be examined: first, that a variably-defined pre-psychotic period (e.g. nonspecific prodrome, CHR/ARMS state, etc…) exists in a large number of patients with psychosis; and second, that these states can be accurately and reliably identified and measured using current methodologies. Our results seem to support the first assumption (i.e. that a large proportion of patients experience a prodrome). Indeed, the observation that disparate methodologies tend to generate similar estimates might increase our confidence in the general finding that the *majority* of patients experience a prodrome. However, the possibility that even a minority (21.7%) of patients experience no prodrome raises questions about measurement approaches, the underlying concepts being appraised and captured, and implications for the structure and function of next-generation services.

### Are there truly patients who do not experience a prodrome?

A clear possibility is that a sizeable minority of patients experience a rapid change from a state of relative wellness to florid psychosis, without an intermediate period of nonspecific or sub- threshold symptoms - akin to previous observations of “acute” (vs. “insidious”) onset of psychosis (McGlashan, 2008; Beiser et al., 1993; Compton et al., 2008; Ito et al., 2015; Morgan et al., 2006). This subgroup has clinical relevance because it is thought to have a better prognosis (Kanahara et al., 2013; McGlashan, 2008), suggesting potentially different neurobiological mechanisms or developmental pathways underlying both their onset of illness, and perhaps the illness itself. Identifying these differences in clinical trajectory and neurobiology may ultimately lead to improved or tailored treatments for this and other subgroups. Additionally, because these patients’ putatively rapid transition to psychosis leaves little opportunity for them to be identified by CHR or general/nonspecific early-intervention services during the prodrome, services would need to be alert to this group and have intake mechanisms geared towards rapid diagnosis, assessment and treatment. A second consideration is that this subgroup *does* in fact experience a prodrome, but simply one that is less easy to measure or that is not captured by the majority of current assessment methodologies, for example because of difficulties in recalling prodromal symptoms (the vast majority of studies considered here relied on retrospective recall or records), or because the prodrome they experience is qualitatively different to the prodrome captured by most current methods.

In summary, it is possible that a sizeable minority of patients do not experience a recognizable prodrome, but it is at least equally plausible that all patients experience some form of prodrome that is difficult to recall, transient, or challenging to identify or measure. Clarity on which of these alternatives is the case (and, should the latter alternative be the case, on what form the prodrome (or prodromes) not reliably measured by current methods takes) would provide critical knowledge to inform the breadth of feasible targets for psychosis prevention. The importance of this question for the structure and function of mental health services is not in doubt (Shah et al., 2022; Brady et al., 2023); however, resolving it requires that the field achieve a consensus definition of the prodrome, operationalizes it in a manner that can be consistently applied, and then generates prospective data from a range of settings which can then be compiled.

### Is the APS definition adequate?

There is currently a great deal of clinical and research effort aimed at determining how to best provide care for, and predict transition to psychosis amongst, patients who meet the criteria for a clinical high-risk state (Fusar-Poli et al., 2012; Addington et al., 2020). The main focus in these settings continues to be on sub-threshold psychotic symptoms, commonly defined by the type and intensity of brief or attenuated positive symptoms present (Fusar-Poli et al., 2013). It is striking, however, to note that the vast majority of the literature on the prodromes experienced by patients who actually develop psychosis do not appear to focus solely on subthreshold positive symptoms of psychosis or APS. Rather, the prodrome has frequently been appreciated as inclusive of a range of affective, negative, positive, non-specific, basic, cognitive, and other symptoms. However, given the more specific definition of the prodrome contained in APS-only studies, it is perhaps surprising that these seem to yield similar estimates of prodrome prevalence, rather than meaningfully lower estimates as one might assume due to their less expansive symptom criteria. In support of this assumption, there are indeed higher prodrome prevalences (close to 100%) when broader definitions (including symptoms beyond the APS definition) are applied to identical datasets (Shah et al, 2017, Cupo et al., 2021). Nonetheless, all three definition subgroups are within 5% of each other’s estimates and lie within each other’s confidence intervals. Such findings suggest that most patients who eventually develop psychosis will at some earlier point experience APS, even if their initial symptoms are nonpsychotic ones (Shah et al., 2017).

If APS are a relatively late-stage symptom cluster, occurring after changes in mood, cognition, social function, and other prodromal symptoms (Hafner et al., 1995; Cupo et al., 2021; Solmi et al., 2021), then are APS-only definitions of prodrome sufficient? With the ultimate objective being to identify patients and intervene early in order to maximize clinical benefit, interventions relying on APS-based definitions may overlook opportunities to identify or delay the onset of psychosis. Indeed, APS-specific interventions may have relatively limited effectiveness even with respect to reducing APS symptoms or transition rates for patients at the CHR stage (Davies et al., 2018; Worthington et al., 2021). As such, while APS may be a necessary and important part of an eventual gold-standard prodrome definition, they may not be adequate, especially when taking heterotypic trajectories into account. Recent work on initiatives such as HiTOP, clinical staging, and p-factor theory (Carrión et al., 2017; Caspi et al., 2014; Kotov et al., 2017; Shah et al, 2020) have all suggested that illness development occurs in a pluripotential and transdiagnostic manner, prompting a better appreciation of the heterotypy inherent in the risk, onset and course of mental illnesses. Our findings, and our recommendations below, are consistent with this understanding of illness development and the need to develop services accordingly.

### Prevalence and Heterogeneity

Because the I2 is a poor measure of heterogeneity in proportion meta-analyses, the use of prediction intervals and subgroup analyses is strongly recommended (Migliavaca et al., 2022). Our prediction interval runs from 41.1% to 93.6%, demonstrating the large heterogeneity in estimates of rates of prodrome between studies in the main analysis. Per subgroup analyses, high heterogeneity (in terms of both I2 and PI) persists even when attempting to group studies by the instruments used, setting, or the methodological approach. While there is some degree of variability across these subgroups (likely due in part to their substantial overlap), the differences between the estimates produced are relatively modest in comparison to the heterogeneity within each subgroup; this along with overlapping confidence intervals makes interpreting these differences challenging.

The fact that the large inconsistency between study estimates persists even when subgrouping studies suggests that the inconsistency is not easily attributable to differences in specific constructs or methodologies but may instead draw on differing conceptualizations or operationalized definitions of the prodrome as well as differing research practices. This should underscore the extent to which improved uniformity of assessment of the prodrome in practice will be critical to obtaining clarity on the question of prodrome prevalence, with corresponding implications for our mechanistic understanding of psychosis onset, treatment development, and service delivery.

### Strengths and Limitations

This is, to our knowledge, the first systematic review of prodrome proportion. Strengths include the incorporation of studies in multiple languages in recognition of the many ways in which the prodrome has been operationalized globally, and the fact that we integrated various definitions of the prodrome while also disaggregating them in subgroup analyses. These subgroup analyses also examined potential sources of heterogeneity. The high I^2^ values reported here are common for prevalence meta-analyses, especially with large numbers of studies, and limit our ability to interpret the I^2^. In this case, we followed the recommendation of Migliavaca et al., 2022 who suggested conducting sensitivity analyses (including subgroup analyses) and reporting prediction intervals to better examine heterogeneity.

The most significant limitation in this review is the substantial heterogeneity across studies. Despite concerted attempts via prodrome category or subgroup analyses, we were unable to identify clear explanations for this. This suggests that the estimates we have identified should be interpreted with caution, and there may be unidentified variables that account for this inconsistency. It also implies that there are meaningful differences between the ways in which different research groups carry out their work that cannot be explained by broad methodological choices, and which are in turn inherently linked to the absence of standard definition of, technique for the measurement of, or lack of consensus in conceptualizing the prodrome. The lack of complementary measures (such as validated biomarkers) which could reduce ambiguity in clinical measurement and therefore potentially improve the reliability of the results presented here, is also a challenge to overcome.

Second, our review yielded few prospective studies in which prodrome definitions could be tested with respect to their predictive validity. Instead, the vast majority of studies identified relied on retrospective definitions of the prodrome, and as such on the recall of patients or the accuracy of medical records not created with the documentation of the prodrome in mind.

Despite our subgroup results suggesting that studies with theoretically shorter recall periods (FEP clinic studies) do not differ meaningfully from those with potentially longer recall periods, some degree of recall bias is likely to remain in the majority of reported proportions.

Researchers may have employed differing skills or effort levels when soliciting retrospective data from patients, families, or medical records - an unmeasured but potential source of heterogeneity. While this may be a key element in the explanation of our results, it can only be accounted for in future prospective studies, as we will discuss below.

Third, we note the lack of data on prodrome prevalence from South America, Africa, and Asia (including the Middle East). Cultural differences in the experience and conceptualization of psychiatric symptoms have long been recognized, which means that the findings from this review may not reflect or be directly generalizable to these jurisdictions. It is, however, reassuring to note that there were no meaningful differences in prodrome prevalence rates by the regions we could include, which does suggest some conserved phenomenology (see the supplementary material).

Fourth, the sample considered here is majority male; while this is not inconsistent with the demographics noted in first episode programs (see Table 1), women tend to have different ages and patterns to psychosis onset (Hafner et al., 1995; Ferrara & Srihari 2020; Brand et al., 2022; Carter et al., 2022; Reicher-Rossler et al., 2018) and so future efforts may need to focus on understanding gender differences in the prodrome as well.

Fifth, we note the lack of differences in prodrome point estimates between the three prodrome definitions. Part of this may be explained by the overlap between groups, and this may limit our ability to interpret this finding. For example, the majority of studies representing the non-specific and specified broad groups would have included APS as part of their definitions. However, it is striking that the non-specific definitions produced similar estimates as those studies with well-operationalized definitions.

Finally, the primary question (and its focus on prodrome prevalence) relied on the prodrome being “absent” or “present”. This may have reduced the resolution of the data available and precluded an analysis of the prodrome as a spectrum of symptoms and severities. This was necessary, however, in order to generate a metric which could be compared between studies, given the inconsistency in definitions between studies.

### Recommendations for the field

The persistent heterogeneity across our analyses leads us to recommend a concerted effort to generate both a consensus definition of the prodrome, as well as a validated and universal mechanism for measuring it. Only through a large-scale, multi-site and multi-country prospective study can it be determined what proportion of individuals who develop psychosis do experience a prior prodrome (and what form or forms this takes). A prospective design with standardized and reproducible assessment methodology can enable a comprehensive range of potential prodromal symptoms to be captured while minimizing variations in researcher efforts and practices.

In addition to standardization and reproducibility, any such study would require broadly scoped, longitudinal, and temporally dense sampling of participants over an extended period of time. While the definition of the prodrome based on symptoms alone has launched and enabled decades of productive research and the development of novel clinical infrastructures, our results suggest that revised definitions of the prodrome may need to be inclusive of additional dimensions beyond symptoms alone in order to have predictive validity, which can then be used to direct services and assist in the development of novel treatments. As such, novel techniques which can be easily implemented at scale should be used to provide augmenting measures which may be clinically meaningful (Shah et al., 2020b; Benrimoh et al., 2022). These would include computerized cognitive batteries (e.g. August et al., 2012), performance on both existing and novel computational tasks (e.g. Powers et al., 2017; Teufel et al., 2015; Kafadar et al, 2022; Vercammen et al., 2010; Benrimoh et al., 2022), and potentially digital phenotyping (Huckvale et al., 2019) and biomarkers (Veronese et al., 2021; Trovão et al., 2019; Mirzakhanian et al., 2014; Fernandes et al., 2020). These extra measures may, for example, help differentiate the cognitive changes seen in a patient with depression from those indicative of an incipient psychosis. Data from this cohort would allow different prodrome definitions to be tested and selected based on a) predictive validity and b) the capacity to differentiate patients who will develop psychosis from those who will develop other mental health conditions.

Such a study would undoubtedly require immense cost and effort, but would nonetheless be worthwhile. Almost by definition, prevention or delay of early psychosis requires an understanding of what proportion of patients experience a prodrome and the form this takes. Ninety years of research - the majority of it retrospective in nature - has been unsuccessful in this endeavor, indicating the need for a concerted and prospective attempt with prospective approaches, inclusive of but not restricted to subthreshold psychotic symptoms.

Crucially, the conduct of this study in a prospective manner as described above would enable the creation not necessarily of a single definition of a unitary ‘prodrome’ but rather of a staged definition (Nieman & McGorry, 2015; Shah et al., 2020a), potentially with subgroups with distinct progression trajectories (including the possibility of a subgroup with no or very short prodromal periods). This staged definition would allow for the development of screening instruments or interventions best suited (in both form and intensity) to a specific stage. As a potential example of this consistent with current conceptualizations, a two stage definition might consist of an “early” prodrome corresponding to nonspecific symptoms, and a “late” prodrome in which subthreshold psychotic symptoms have emerged (Keshavan et al., 2011). The addition of novel measurement modalities (e.g. biomarkers, computational tests) may also add to the staging model as some of these measures may change in concert with, or ideally predict, changes in stage and these may in turn contribute to both screening efforts and treatment targets in the future. Overall, having a valid definition would allow us to improve screening processes, and potentially to introduce screening at scale. It would allow us to finally determine if there are some patients who will simply not develop a prodrome, and to plan services accordingly. Most importantly, it would allow further mechanistic research on individuals in “true” prodromal states (however defined) which, in turn, could allow us to develop novel treatments that could delay- or perhaps prevent- the onset of psychosis.

## CONCLUSIONS

In this systematic review and meta-analysis, we present for the first time an estimate of the prevalence of prodrome prior to psychosis onset across nearly 90 years of research. Our estimate of 78.3%, while comprehensive, reveals a high degree of heterogeneity which largely remained even when subgrouping studies based on definitions of prodrome or on methodologies, and which was associated with a wide predictive interval. We argue that a way forward is a large-scale, prospective, densely sampled cohort study using both rigorous symptom assessments and cognitive, behavioral, and computational batteries that could generate gold-standard prodrome definition(s). The findings of such a study would serve to focus world-wide efforts to delay or prevent the onset of psychosis.

## Supporting information

SM: Extraction Form

PRISMA Abstract Checklist

PRISMA Checklist

Table 1: Included Studies

Table 2: Prodrome Definitions

Table 3: Subgroup Analyses

## Data Availability

DATA AND CODE AVAILABILITY: Analyses were carried out using commercially available software. Data required to reproduce analyses are included in Table 1 and 2.

## ACKNOWLEDGEMENTS

We would like to extend our sincere thanks to the library sciences department at Yale University for their significant contributions to the process of finding, requesting, and making available articles for this review. We would also like to thank all those authors who made a significant effort to provide us with data missing from their articles in support of this review. These included: Dr. Jackson, Dr. Riecher-Rossler, Dr. Yildizhan, Dr. Pelizza, Dr. Ehrenreich, Dr. Barrantes, Dr. Velthorst, Dr. Salvatore, Dr. Dazzan, Dr. Conus, Dr. McGorry, Dr. Skokou, Dr. Woodberry, Dr. Van Os, Dr. Sullivan, Dr. Mustonen, and Dr. Renwick.

## FUNDING AND CONFLICTS OF INTEREST

No funding was received in connection to this review. D.B. is a shareholder, founder, and employee of Aifred Health, a digital mental health company which was not involved in any way with this review and whose work is not relevant to the aims of this review. D.B. did not receive funding from Aifred Health in connection to this review.

## DATA AND CODE AVAILABILITY

Analyses were carried out using commercially available software. Data required to reproduce analyses are included in Table 1 and 2.

## SUPPLEMENTARY MATERIAL

### Prospero listing

David Benrimoh, Abigail Wright, Melissa Funaro, Peter Phalen, Viktor Dlugunovych, Vinod Srihari, Scott Woods, Keith Gallagher, Albert Powers, Maria Ferrara, Jai Shah. A Systematic Review of Prevalence of the Prodrome Preceding First Episode Psychosis. PROSPERO 2021 CRD42021239797 Available from: https://www.crd.york.ac.uk/prospero/display_record.php?ID=CRD42021239797

The only main change from the listed protocol was the inclusion of proportion meta-analysis and predictive interval; these were added once it was clear sufficient data would be available to support these analyses.

### Supplementary Section 1: Supplemental methods and search strategies

The search was intentionally broad and expected to produce many false positives. This was done in order to capture studies which included estimated of prodrome prevalence even if this was not the study’s main endpoint.

While studies of patients with solely drug induced psychosis were excluded, we allowed studies in which a subset of the study sample experienced drug induced psychosis, as these subsets were generally a small proportion of the overall study sample, and this allowed the inclusion of several important studies.

We excluded conference abstracts because these do not generally contain sufficient details about methodology required by the objectives of this review.

In our subgroup analyses, we compared the inconsistency and prevalence estimates from studies which included two approaches to measuring prodrome: those which utilized as part of their definition the DSM3 prodrome definition, which is syndromal in nature; and those which were inspired by the IRAOS instrument, which focuses on a more symptom-driven view of the prodrome. The intent was to determine the differences between syndrome and symptom based approaches, and the extent of inconsistency even *within* a family of studies using similar references. Finally, we considered studies which only had patients with schizophrenia spectrum disorders, and compared them to studies which had more heterogeneous populations (e.g. affective psychoses, delusional disorders, substance-induced psychoses).

### All Search Strategies

Ovid MEDLINE(R) ALL <1946 to March 02, 2021>

|  |  |
| --- | --- |
| 1 | Schizophrenia/ or Psychotic Disorders/ or (psychos?s or psychotic or schizo*).tw,kw. |
| 2 | (first admission or first admit* or first episod* or first hospital* or first present* or onset or preced* or pre-episod* or prepsycho* or preepisod* or pre-psycho* or early schiz*).tw,kw. |
| 3 | ((early or first) adj (psychoses or psychosis or psychotic or schizo*)).tw,kw. |
| 4 | 2 or 3 |
| 5 | Prodromal Symptoms/ or (((insidious or acute or nonacute or non acute) adj onset) or prodrom* or "at risk mental state*" or ARMS or clinical high risk or CHR or CHR-P or ultra high risk or UHR or subthreshold psycho* or cognitive symptom* or basic symptom*).tw,kw. |
| 6 | 1 and 4 and 5 |

Embase <1974 to 2021 March 02> (Ovid)

|  |  |
| --- | --- |
| 1 | schizophrenia/ or psychosis/ or (psychos?s or psychotic or schizo*).tw,kw. |
| 2 | (first admission or first admit* or first episod* or first hospital* or first present* or onset or preced* or pre-episod* or prepsycho* or preepisod* or pre-psycho* or early schiz*).tw,kw. |
| 3 | ((early or first) adj (psychoses or psychosis or psychotic or schizo*)).tw,kw. |
| 4 | 2 or 3 |
| 5 | prodromal symptom/ or (((insidious or acute or nonacute or non acute) adj onset) or prodrom* or "at risk mental state*" or ARMS or clinical high risk or CHR or CHR-P or ultra high risk or UHR or subthreshold psycho* or cognitive symptom* or basic symptom*).tw,kw. |
| 6 | 1 and 4 and 5 |
| 7 | 6 not ((exp animal/ or nonhuman/) not exp human/) |

APA PsycInfo/PsycBooks <1806 to February Week 4 2021> (Ovid)

|  |  |
| --- | --- |
| 1 | Schizophrenia/ or psychosis/ or (psychos?s or psychotic or schizo*).mp. |
| 2 | "Onset (Disorders)"/ or (first admission or first admit* or first episod* or first hospital* or first present* or onset or preced* or pre-episod* or prepsycho* or preepisod* or pre-psycho* or early schiz*).mp. |
| 3 | ((early or first) adj (psychoses or psychosis or psychotic or schizo*)).mp. |
| 4 | 2 or 3 |
| 5 | prodrome/ or (((insidious or acute or nonacute or non acute) adj onset) or prodrom* or "at risk mental state*" or ARMS or clinical high risk or CHR or CHR-P or ultra high risk or UHR or subthreshold psycho* or cognitive symptom* or basic symptom*).mp. |
| 6 | 1 and 4 and 5 |
| 7 | 6 not (Animals not (Animals and Humans)).mh. |

Web of Science Core Collection (Clarivate)

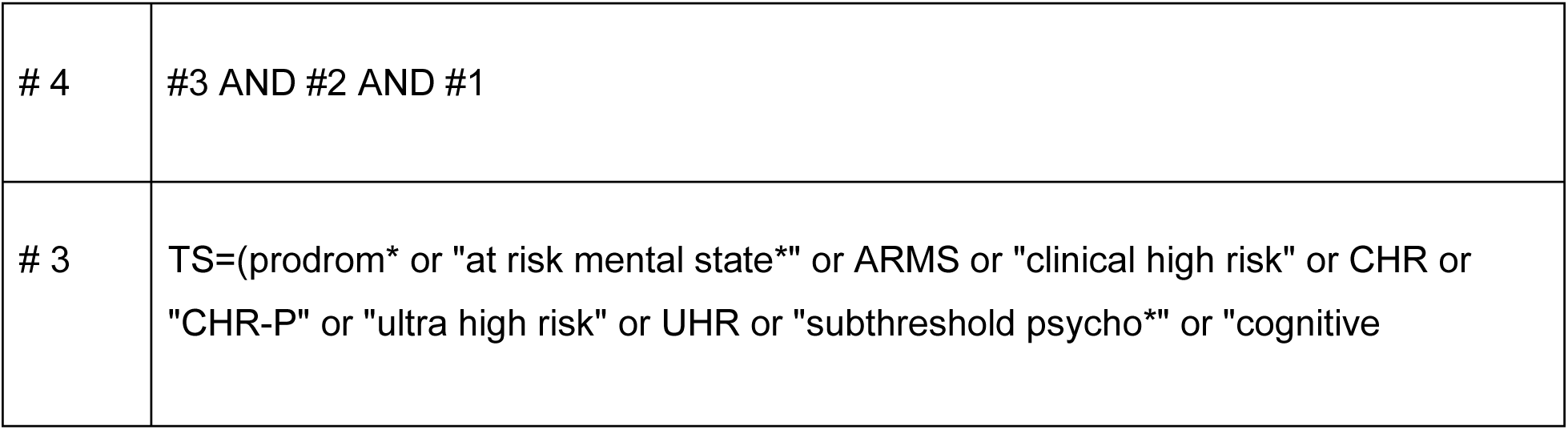
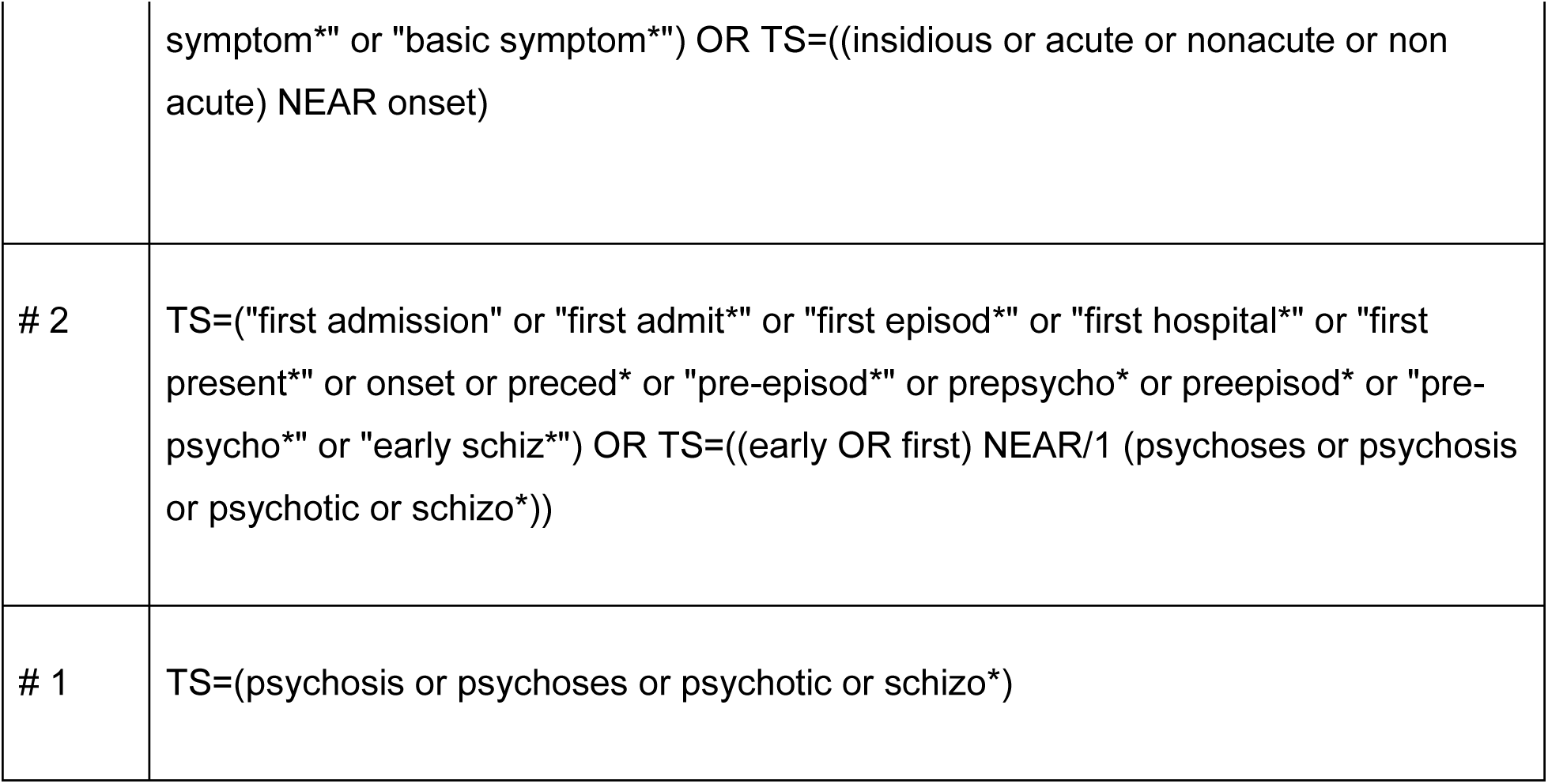

|  |  |
| --- | --- |
| # 4 | #3 AND #2 AND #1 |
| # 3 | TS=(prodrom* or "at risk mental state*" or ARMS or "clinical high risk" or CHR or "CHR-P" or "ultra high risk" or UHR or "subthreshold psycho*" or "cognitive |
|  | symptom*" or "basic symptom*") OR TS=((insidious or acute or nonacute or non acute) NEAR onset) |
| # 2 | TS=("first admission" or "first admit*" or "first episod*" or "first hospital*" or "first present*" or onset or preced* or "pre-episod*" or prepsycho* or preepisod* or "pre-psycho*" or "early schiz*") OR TS=((early OR first) NEAR/1 (psychoses or psychosis or psychotic or schizo*)) |
| # 1 | TS=(psychosis or psychoses or psychotic or schizo*) |

Cochrane

|  |  |
| --- | --- |
| #1 | (psychosis or psychoses or psychotic or schizo*):ti,ab,kw |
| #2 | ("first admission" or "first admit*" or "first episod*" or "first hospital*" or "first present*" or onset or preced* or "pre-episod*" or prepsycho* or preepisod* or "pre-psycho*" or "early schiz*"):ti,ab,kw OR ((early OR first) NEAR/1 (psychoses or psychosis or psychotic or schizo*)):ti,ab,kw |
| #3 | ((prodrom* or "at risk mental state*" or ARMS or "clinical high risk" or CHR or "CHR-P" or "ultra high risk" or UHR or "subthreshold psycho*" or "cognitive symptom*" or "basic symptom*") OR TS=((insidious or acute or nonacute or non acute) NEAR onset)):ti,ab,kw |
| #4 | #1 AND #2 AND #3 |

### Supplementary methods: Data Extraction Form

Attempts were made during extraction to determine the recall interval for patients in retrospective studies (e.g. the time between interview and symptom onset) but this could not be extracted reliably from enough studies for us to be confident in using this in an analysis. The extraction form is included in a PDF attachment. Any data fields included in the form which are not included in Table 1 and 2 were excluded because data was systematically missing from studies or because it was decided by the authors not be useful in the context of the objectives of the review. No data was imputed.

#### Further comments on a potential prospective study

The ideal population for a prospective study aimed at determining and validating a staged prodrome definition would be a set of help-seeking catchment-based samples; to make these samples practical, they could be limited to children, adolescents, and adults up until age 35 (to align with admission criteria for some early intervention services who seek help for *any* mental health reason). The focus on a help-seeking population would likely bias the sample against those with acute onset or no prodromal symptoms; as such a rigorous case-finding process would need to be put in place in each catchment area to identify patients who develop psychosis and who are not already being followed in the prospective study, and general population sampling methods could also be used in order to supplement help-seeking based recruitment. Detailed retrospective data, validated by multiple sources (e.g. family or school report), would also need to be captured from every patient entering the study, to account for symptoms that may have been experienced prior to study entry; however, rigorous training and frequent inter-rater reliability testing will be required to mitigate the effects of recall bias. Another possibility for a patient sampling method would be a multi-country birth cohort, with entry of participants at age 10; further feasibility studies would be required in order to determine if the temporally dense sampling we recommend would be feasible in this type of study.

#### Analysis by definition, country and publication year

Further analyses were conducted to assess the impact of changing definitions over time and regional definitional traditions on prevalence rates and are presented in the supplementary material. Using the three prodrome definition categories above, we conducted an ANOVA to determine whether the prevalence rates reported were significantly different across groups. ANOVA was also used to determine if broad geographical areas (continents) differed in terms of prodrome prevalence. Correlation analysis was used to determine if year of publication was significantly associated with reported prevalence estimates. As these were secondary analyses, we did not correct for multiple comparisons.

ANOVA demonstrated no significant differences between the three definition groups (df=2, F= 0.1, *p*=0.904). These were also not different using the Q statistic (Q(2)=0.228, p = 0.892) using a meta-analytic approach.

ANOVA testing for differences in reported prevalence rates by region were not significant (df=4, F=0.14, *p*=0.97), see Supplementary Figure 1a. Using meta-regression, no differences were found (see supplementary Figure 1b); this is reinforced by observing the scatterplot (supplementary Figure 1c)

Correlation analyses examining prevalence rate by year of publication were also nonsignificant (Pearson correlation=0.23, *p*=0.104). Meta-regression did demonstrate significance for year (with later years predicting higher rates) but the model was poor and had an R^2^ value of 0; see supplementary figure 2a for model outputs and 2b for scatterplot by year; the relationship observed may simply have been a result of a very low estimate by the oldest study. Visual inspection of the scatterplot does not provide any indication of a meaningful relationship with year. Indeed, when removing the outlier from 1932 with the lowest prevalence rate (22%), the model is no longer significant (see supplementary figures 2c and 2d).

**Supplementary Table 1:**
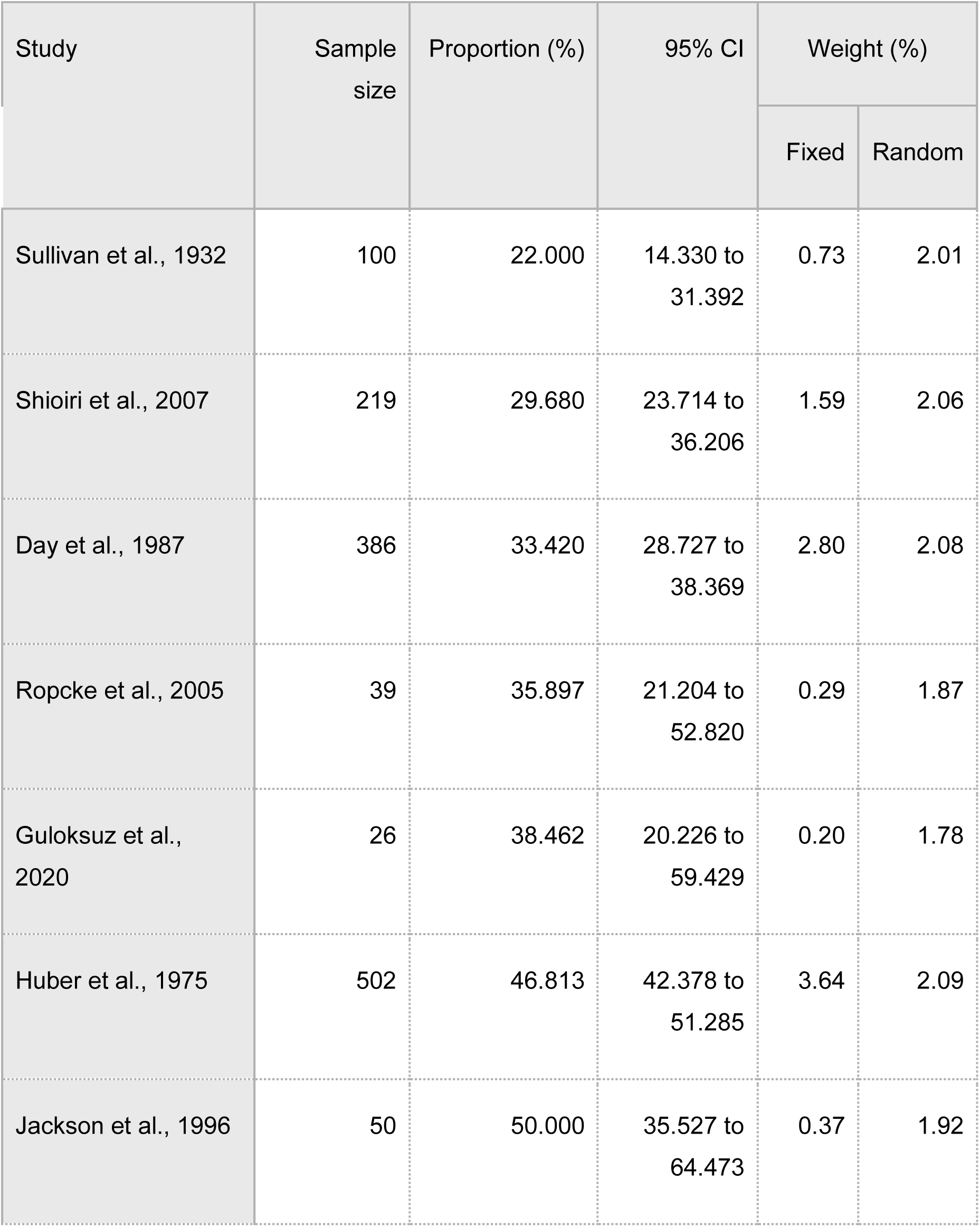
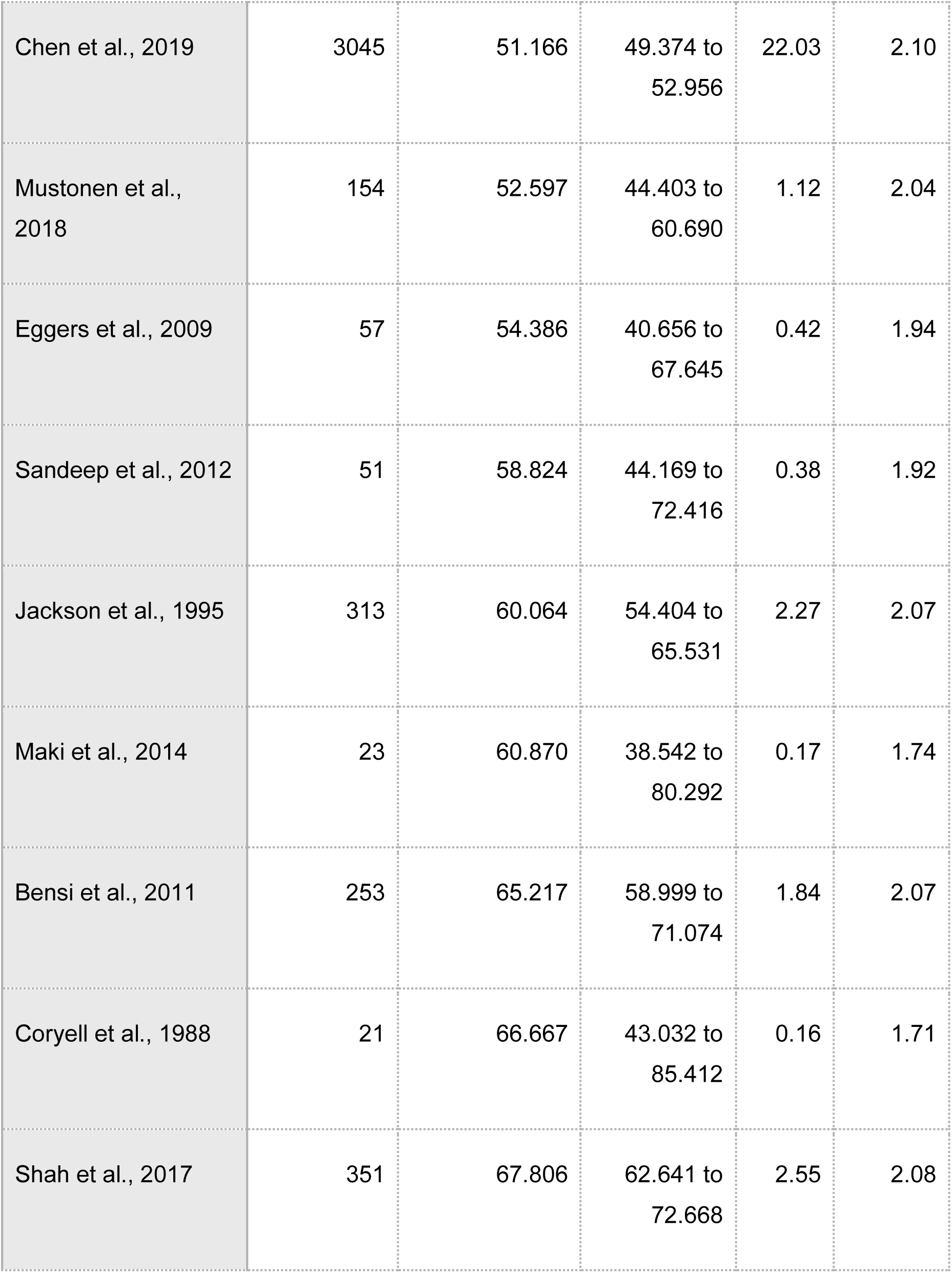
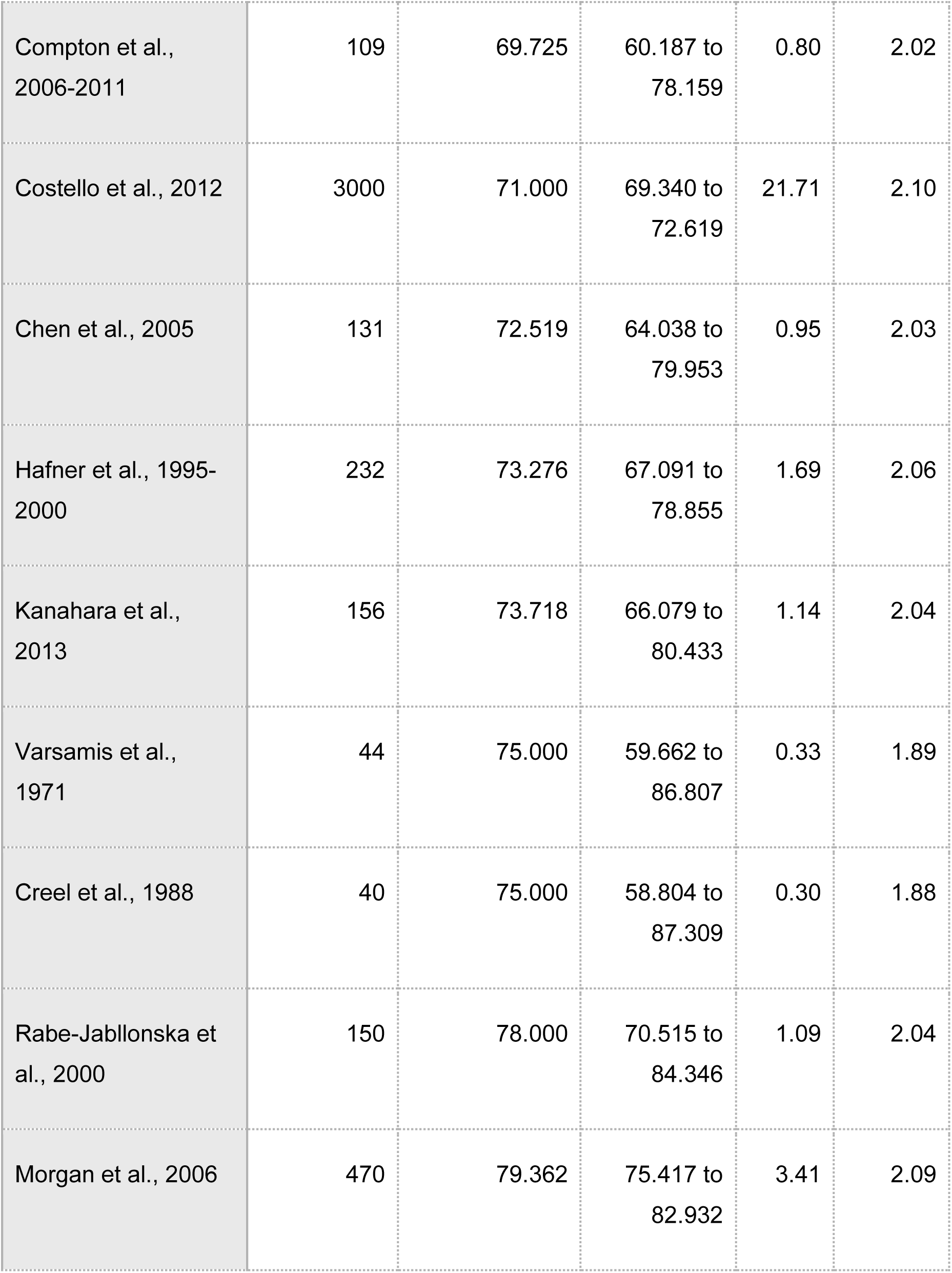
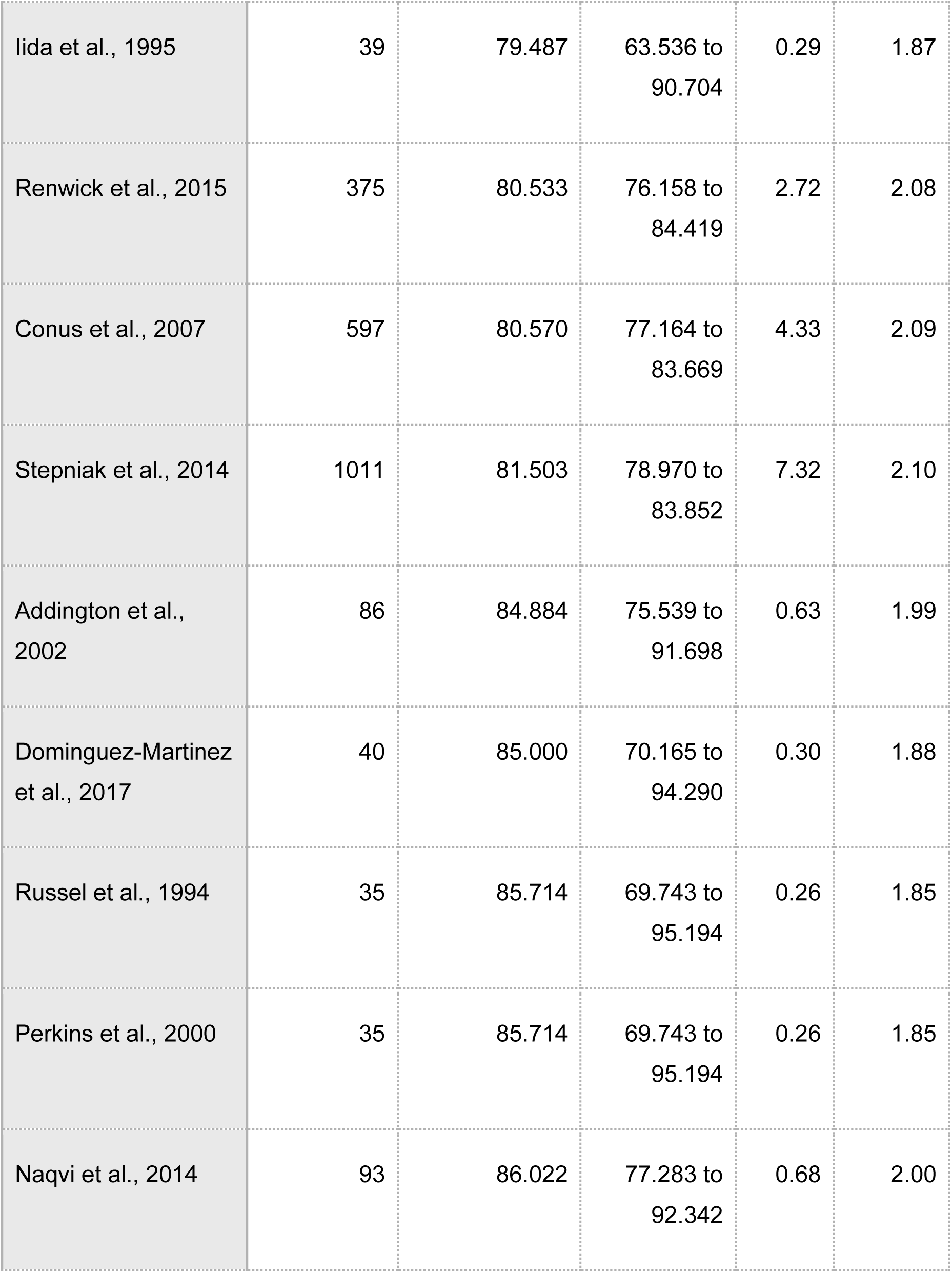
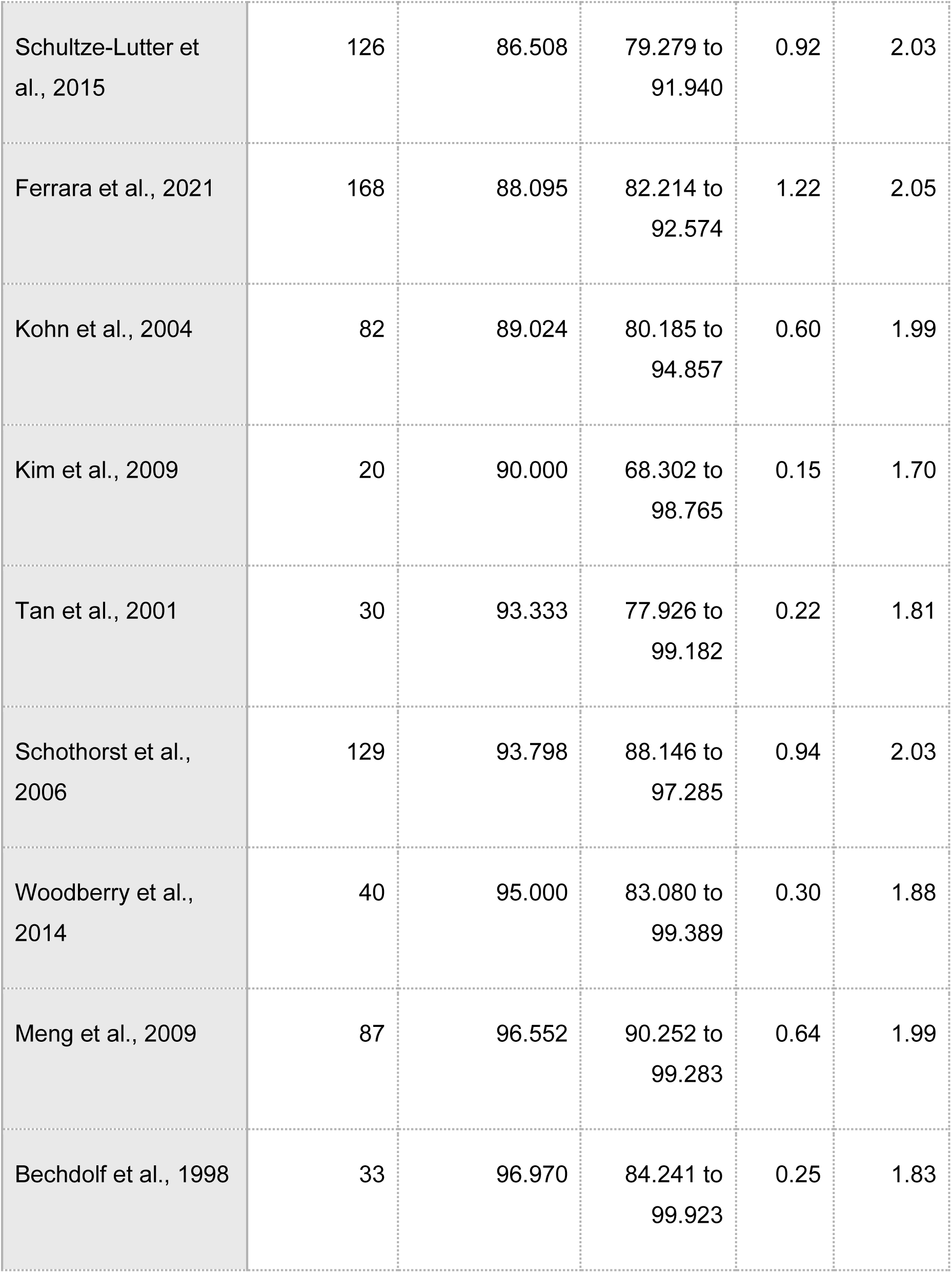
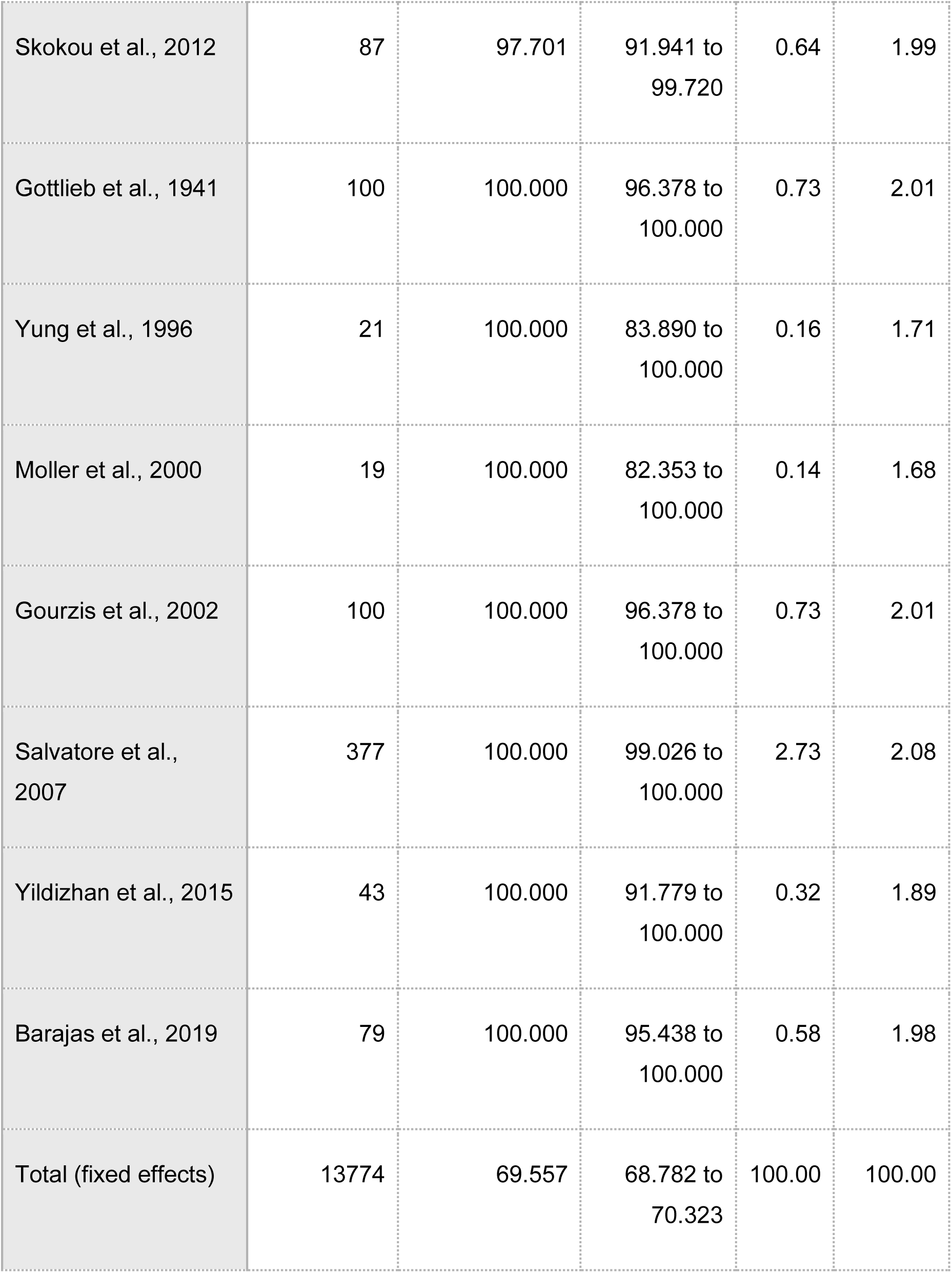
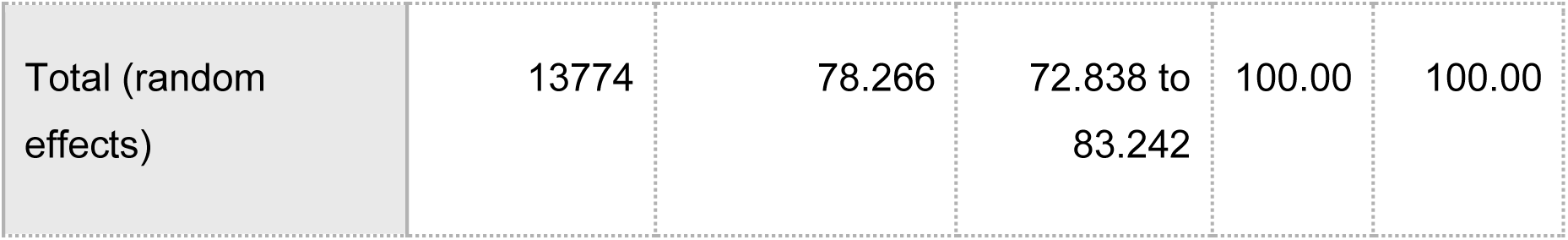
Weights and Estimates for All Studies

**Supplementary Figure 1a:**
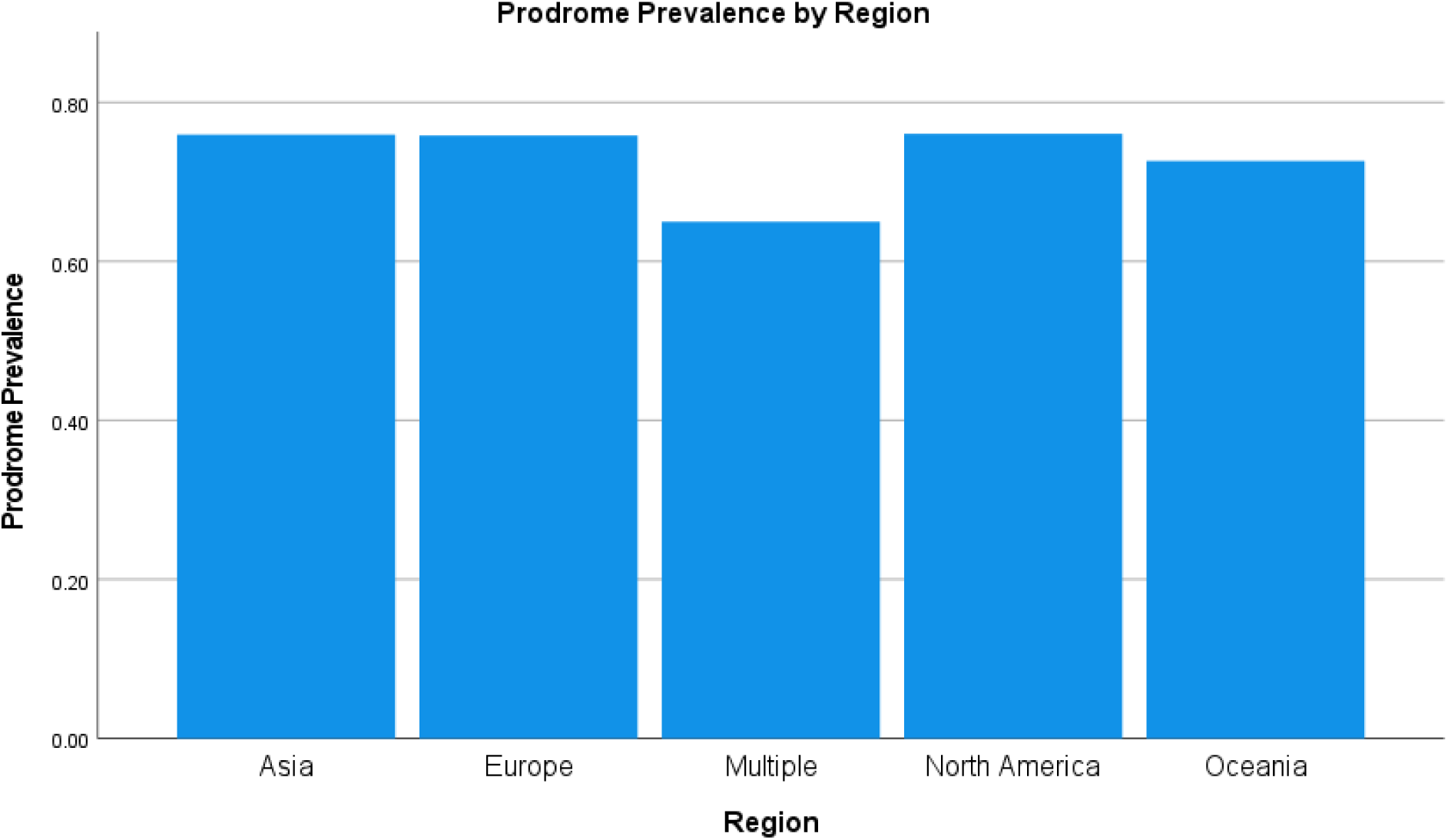
Prodrome Prevalence by Region (mean at the study level)

**Supplementary figure 1b:**
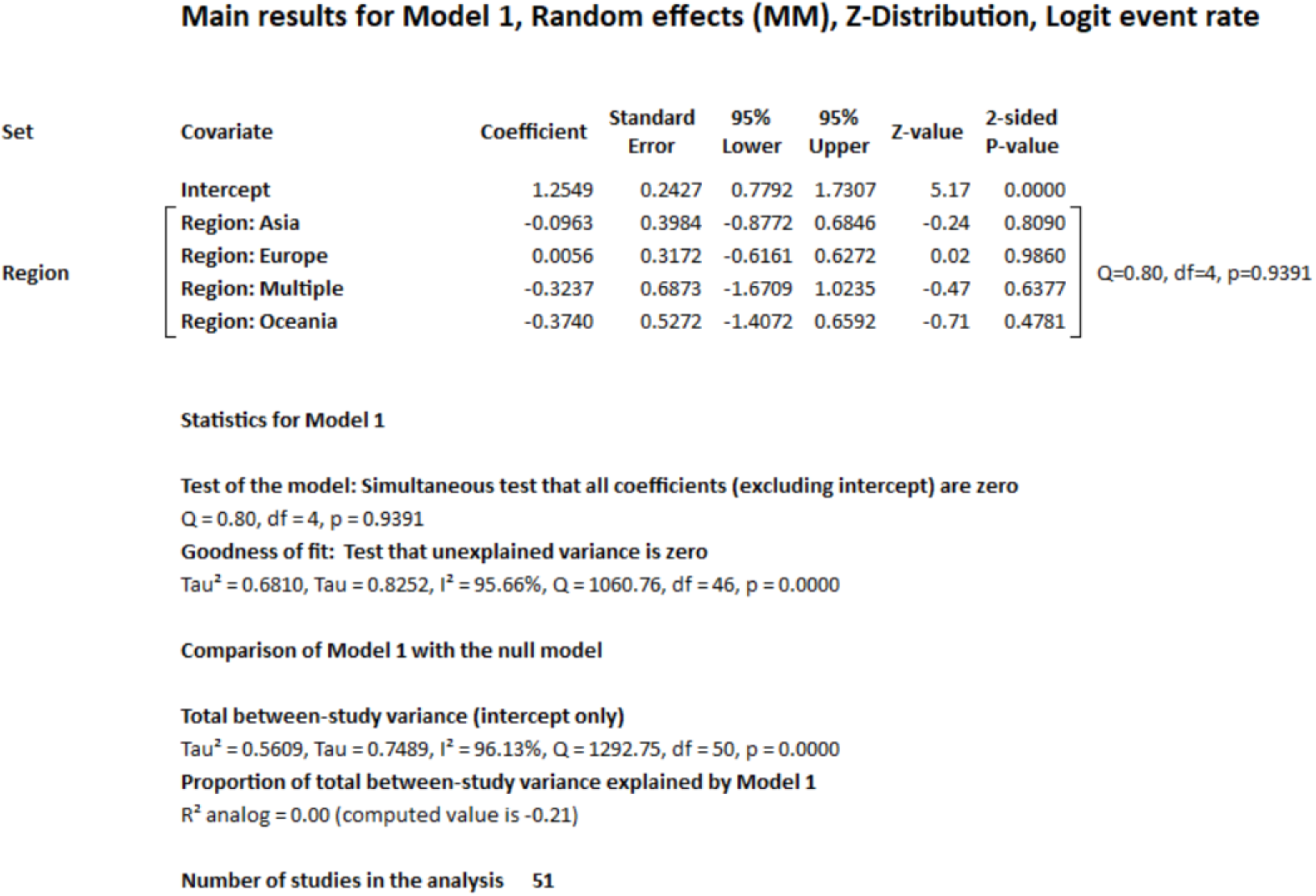
Model Output for Meta-Regression by region (reference region is North America):

**Supplementary figure 1c:**
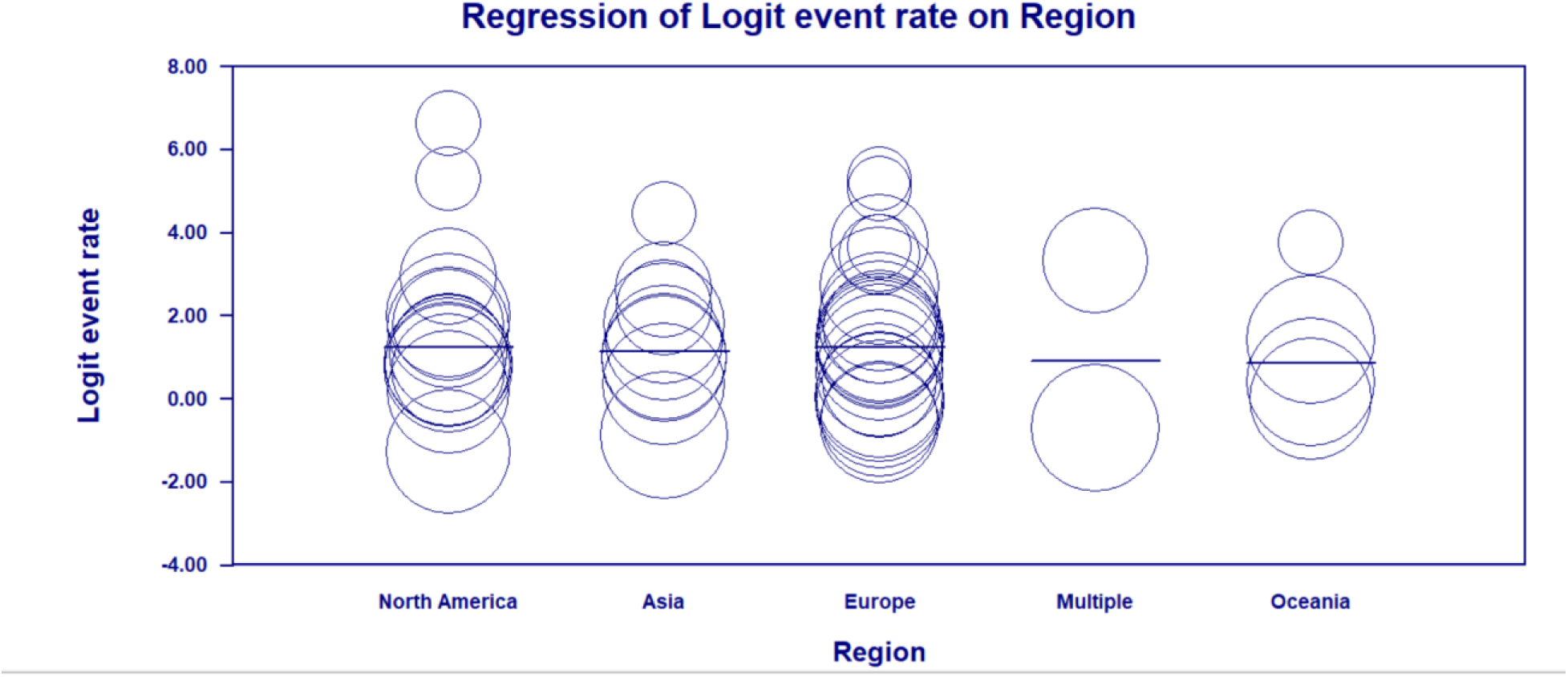
Scatterplot by region:

**Supplementary figure 2a:**
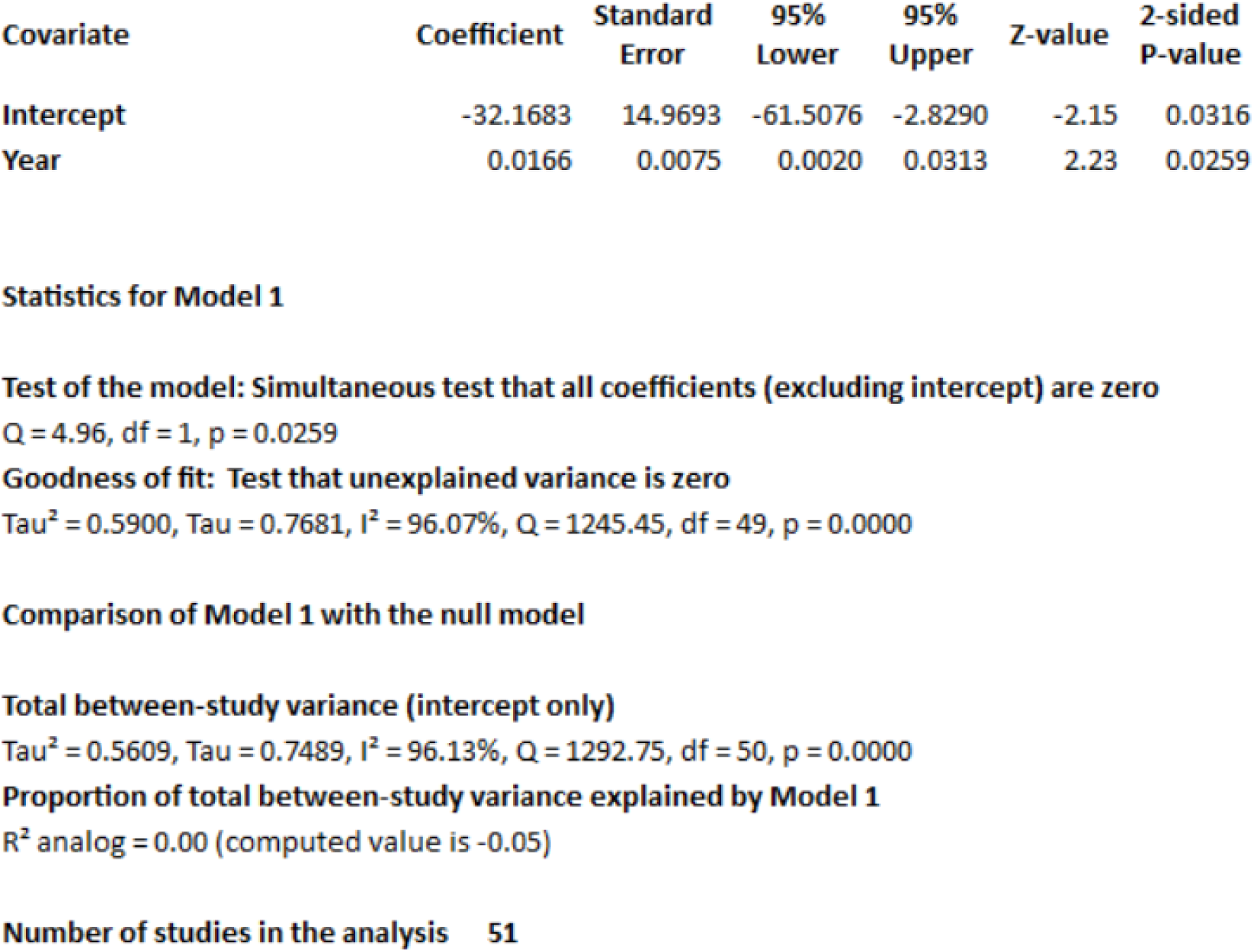
Model Output for Meta-Regression by year:

**Supplementary Figure 2b:**
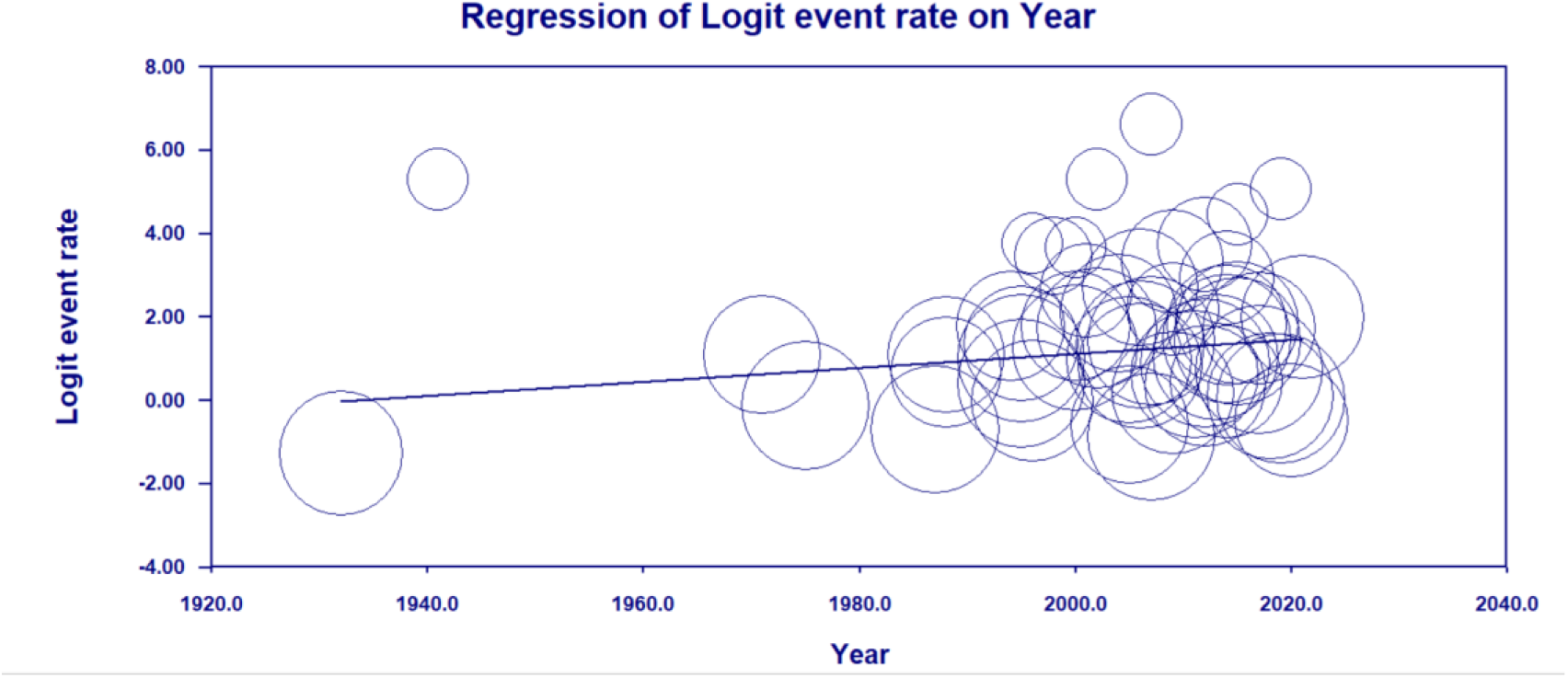
Scatterplot by year

**Supplementary figure 2c:**
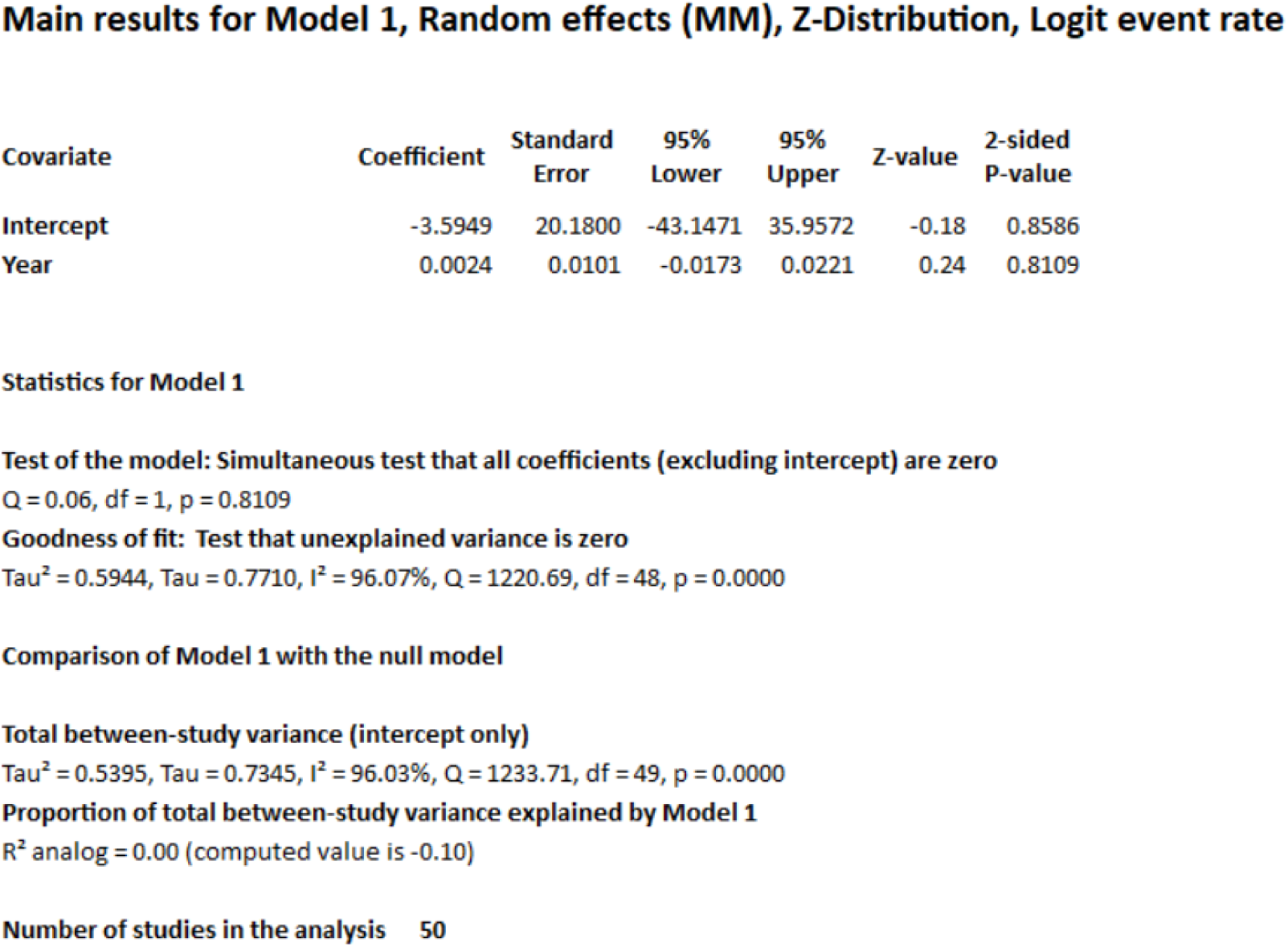
Model output for metaregression by year, removing the outlier

**Supplementary Figure 2b:**
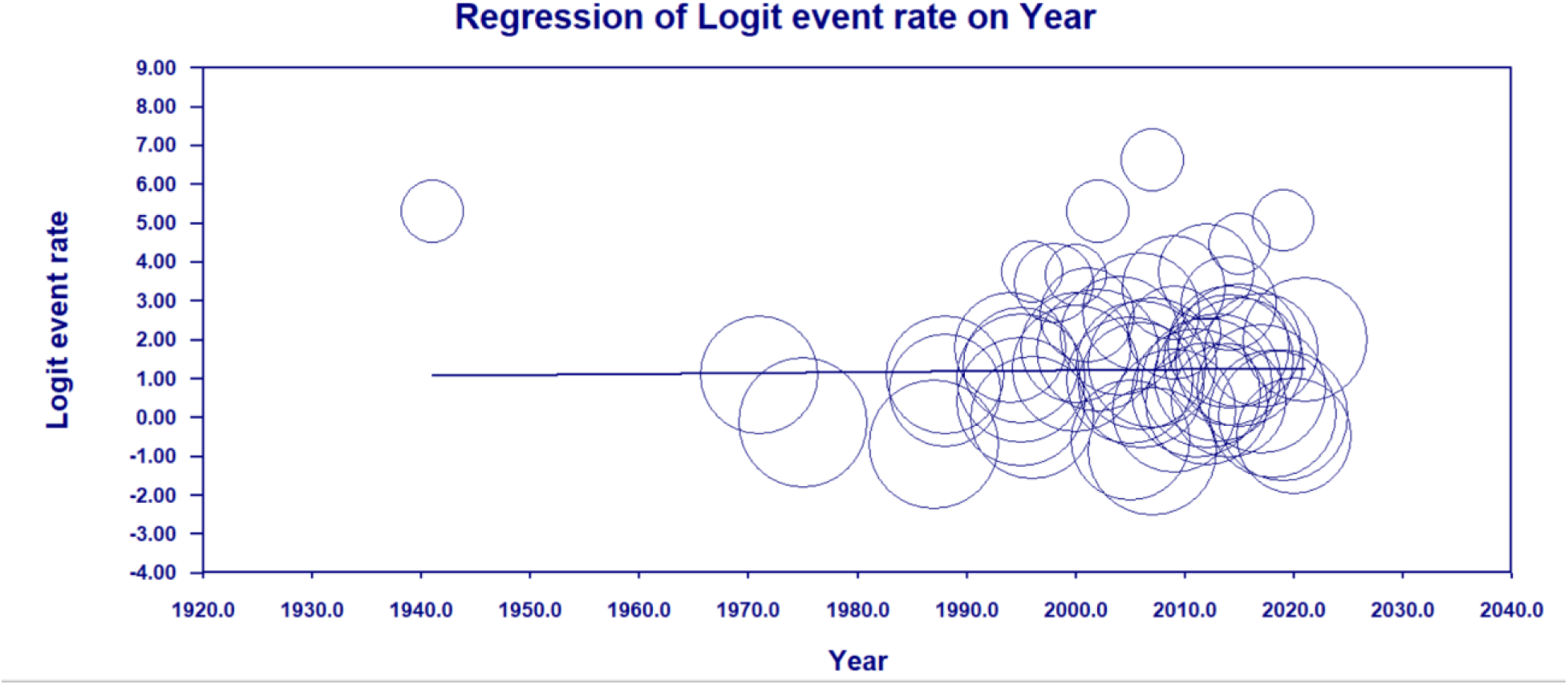
Scatterplot by year, removing the outlier

