## Supplementary material for "On the Proportion of Patients Who Experience a Prodrome Prior to Psychosis Onset - A Systematic Review and Meta-analysis": SM: Extraction Form

### Extraction Form for FEP Review

Please type "NR" if the answer to a question is not reported

**** Switch account

Not shared

Name

Your answer

Are you:

- ○ Reviewer 1
- ○ Reviewer 2 (validation)

Article Title

Your answer

Covidence Number

Your answer

Publication type

- ◯ Original article
- ◯ Thesis
- ◯ Other:

Funding source

- ☐ No funding (explicit statement that there was no funding)
- ☐ No funding reported / absence of comment on funding source
- ☐ Public granting agency (e.g. CIHR, NIH)
- ☐ Foundation
- ☐ Advocacy group/consumer group
- ☐ Other:

Possible conflicts of interest

- ☐ COI reported on, not present
- ☐ COI reported on, present for at least some authors
- ☐ COI not reported on
- ☐ Other:

Study type

- ◯ Prospective cohort study
- ◯ Retrospective analysis/follow back studies
- ◯ Prospective Intervention study
- ◯ National registry study
- ◯ Cross-sectional study
- ◯ Retrospective intervention study
- ◯ Other:

If an intervention study, what was the intervention?

Your answer

Sample size

Your answer

Inclusion criteria of the study

Your answer

Exclusion Criteria of the study

Your answer

Male/female numbers

Your answer

Age, age range, and SD

Your answer

Diagnoses (if multiple, write n for each, e.g. schizophrenia 25, schizoaffective 5)

Your answer

Race/ethnicity (if not reported, state not reported)

Your answer

What was the recruitment method? (e.g., telephone, flyers, clinician recruitment)

Your answer

Describe the sampling method (e.g. simple random sample, convenience)

Your answer

Describe the study population?

Your answer

Country or Countries in which the study was conducted (where data was actually collected)

Your answer

Study site(s) (e.g. McGill University vs. Local Health Clinic A) ; for studies at multiple (more than 3) institutions, note multicenter study and we will go back to it if we need

Your answer

Study setting(s) (e.g. rural, university center etc...)

Your answer

Years of data collection (year or range)

- ◯ Not available
- ◯ Other:

What length of time did participants spend in the study itself (i.e. the time they were observed for; if the study was cross sectional say "cross sectional")

Your answer

Prodrome definition used by the authors (if an instrument, quote the instrument and the cutoffs; if some other definition, copy paste here).Please comment on the expected time course of prodromal symptoms

Your answer

Was the definition diagnostic/syndromal (e.g., CHR/UHR/ARMS) or symptom-based/dimensional (e.g., hallucinations, depression, cognitive symptoms, withdrawal, “disturbance of perceptions of self”)?

Your answer

Was the definition defined a priori (e.g. defined based on a previously validated definition) or a posteriori (e.g. defined prodrome based on factor analyses of the current sample, or performed an atheoretical tally of reported pre-psychosis experiences)

Your answer

How was the presence or absence of prodrome assessed (i.e. chart review, interview, self-report)

- [ ] Chart review
- [ ] Interview (unstructured)
- [ ] Self report
- [ ] Structured or semi-structured interview of patient with a scale
- [ ] Caregiver/family report
- [ ] Structured or semi-structured interview of patient without a scale
- [ ] Other:

Who assessed the prodrome based on the definition?

- ○ cutoff based on self report scale
- ○ cutoff based on clinician report scale
- ○ chart review by one experienced rater
- ○ chart review by more than one experienced rater
- ○ the impression of an experienced or trained clinician or rater
- ○ the impression of an untrained clinician or rater, or one without significant experience
- ○ Other:

Instruments used to measure the duration, severity, or presence/absence of the prodrome (type "NA" if there was no instrument)

Your answer

mean and standard deviation for continuous prodrome scales (and identify the scale)

Your answer

Was prodrome identified prospectively or retrospectively?

- Prospectively
- Retrospectively
- Other:

Recall interval for prodrome for retrospective studies (how far back did we ask ppl to remember)

Your answer

Did symptoms need to be continuous into the first episode of psychosis or were these any symptoms prior to the first episode ?

Your answer

Information on observed prodromal symptom timecourse, if any; include descriptions of order of symptom appearance

Your answer

Conversion rate to psychosis in general and after prodrome in particular (if a prospective population cohort)

Your answer

Length of prodrome (mean and SD)

Your answer

Number of people with the prodrome

Your answer

“proportion of individuals with a prodrome”, notwithstanding the definition, reported as a percentage

Your answer

Was a primary purpose of the study specifically to identify prodrome prevalence?

- Yes
- No
- Other:

What was the analysis of interest in the study (and what was the purpose of the study)

Your answer

Instruments used to measure the duration, severity, or presence/absence of psychosis

Your answer

Length of psychotic illness if applicable (mean and SD)

Your answer

Other potential sources of bias (including recall bias)/Potential biases which come from the inclusion and exclusion criteria or from recruitment method (from paper limitations or from your own critical review; try and separate the two)

Your answer

Potential methodological issues, from limitations or your assessment, separate the two)

Your answer

key conclusions of study authors relevant to this review, if any

Your answer

Any other comments?

Your answer

Was the sample frame appropriate to address the target population? (JBI guide: https://jbi.global/sites/default/files/2020-08/Checklist_for_Prevalence_Studies.pdf)

- Yes
- No
- Unclear
- Not applicable

Were study participants sampled in an appropriate way?

- Yes
- No
- Unclear
- Not applicable

Was the sample size adequate?

- Yes
- No
- Unclear
- Not applicable

Were the study subjects and the setting described in detail?

- Yes
- No
- Unclear
- Not applicable

Was the data analysis conducted with sufficient coverage of the identified sample?

- Yes
- No
- Unclear
- Not applicable

Were valid methods used for the identification of the condition?

- Yes
- No
- Unclear
- Not applicable

Was the condition measured in a standard, reliable way for all participants?

- Yes
- No
- Unclear
- Not applicable

Was there appropriate statistical analysis?

- Yes
- No
- Unclear
- Not applicable

Was the response rate adequate, and if not, was the low response rate managed appropriately?

- Yes
- No
- Unclear
- Not applicable

Overall quality with respect to our review objectives

- Poor
- Fair
- Good

Do you think this paper should be excluded at this stage?

- Yes
- No

Why do you think this paper should be excluded at this stage?

Your answer

If you are reviewer 2, did you have any discrepancy with reviewer 1 (ignoring individual JBI items)

- Yes
- No
- No discrepancy, just added some detail
- Other:

Submit

Clear form

Never submit passwords through Google Forms.

This content is neither created nor endorsed by Google. Report Abuse - Terms of Service - Privacy Policy

Google Forms
