## Supplementary material for "On the Proportion of Patients Who Experience a Prodrome Prior to Psychosis Onset - A Systematic Review and Meta-analysis": Table 1: Included Studies

| Authors | Country (s) where was conducted / data collected | Years of data collection | Inclusion Criteria | Exclusion Criteria | Recruitment method | Overall quality | Recognized prodrome scale | Study setting | Sample size- | Male number/% | Mean age and (standard deviation) | Prodrome  % |
| --- | --- | --- | --- | --- | --- | --- | --- | --- | --- | --- | --- | --- |
| Sandeep (2012) | India | 2010-2011 | Patents 18-60 with psychotic mania | Comorbid psychiatric or significant medical or neurological illness | Admitted patients | Fair | Y | University health center | 51 | 39/76 | 24.62 | 58.82 |
| Yung & McGorry (1996) | Australia | 1993 | FEP aged 16-30 | Non-psychiatric cause of psychosis, intellectual disability, active psychosis during data collection | Patients recruited from a FEP clinical service | Fair | N | University-affiliated FEP program | 21 | 14/67 | 23.1 | 100 |
| Yildizhan et al. (2015) | Turkey | 2011-2012 | FEP, age </= 20 | Psychotic disorder due to substance use, severe congenital cognitive difficulty preventing interview, previous adequate treatment (equivalent of 6 mg haloperidol/day for 6 weeks) | Admitted patients | Fair | Y | University health center | 43 | 32/74 | 17.38 | 100 |
| Woodberry et al. (2014) | USA | NR | FEP, age 13-45, SZ spectrum | Sensory-motor handicaps, neurological disorders, medical illnesses that significantly impair neurocognitive  function, intellectual disability, education less than 5th grade if under 18 or less than 9th  grade if 18 or older, substance abuse in the past month, substance dependence, excluding nicotine, in the past 3 months, current suicide risk, history of electroconvulsive therapy within the prior 5 years | Recruited from area hospitals, outpatient treatment settings, and the metropolitan Boston community through advertisements, formal outreach presentations, and word of mouth | Poor | Y | University health centers, community and outpatient centers | 40 | 27/68 | 21.9 | 95 |
| Tan &Ang (2001) | Singapore | 1997-1999 | Military servicemen with FEP | Clear-cut non-psychiatric etiology for psychotic symptoms, patients with attenuated or questionable psychotic symptoms for which a firm diagnosis could not be made. | Recruited military personnel presenting with FEP to a military psychological center | Fair | N | Military psychological care center | 30 | 30/100 | 20.6 | 93.33 |
| Sullivan (1932) | USA | NR | SZ, male | Inadequate coupled with failure of the physician to establish satisfactory contact with the patient, intellectual disability coupled with inadequate information, questioning of the schizophrenic nature of the illness | Review of notes from the first 155 cases of SZ admitted to the study center | Poor | N | Urban hospital | 100 | 100/100 | NR | 22 |
| Skokou et al. (2012) | Spain | 2005-2008 | Paranoid SZ, </= 3 episodes | Subjects with non-psychiatric causes of psychiatric symptoms, or psychotic disorder due to substance use or general medical condition | Admitted patients | Fair | Y | University health center | 87 | 54/62 | 30.71 ± 8.68, with a range from 17 to 59 years for males, and 36.47 ± 10.59, with a range from 21 to 65 years for females | 97.7 |
| Shioiri et al. (2007) | Japan | 1999-2004 | SZ | NR | Admitted patients, chart review based on treatment records including reports by family | Fair | N | University health center | 219 | 98/45 | 33.9 | 29.68 |
| Schultze-Lutter et al. (2015) ^a^ | Germany | NR | FEP | See GRNS | Admitted patients | Good | Y | Multicenter inpatient | 126 | NR | 30.1 | 86.51 |
| Schothorst et al. (2006) ^b^ | Netherlands | 1984-2000 | Aged 12-18 with diagnosis of a psychotic disorder | NR | Chart review, outpatients, inpatients, day clinic | Poor | N | University health center | 129 | 86/67 | 16.5 | 93.8 |
| Stepniak et al. (2014) | Germany | NR | SZ and SZ-A | NR | Recruited as part of another study (GRAS) | Fair | N | Multicenter (GRAS) | 1011 | NR | NR | 81.5 |
| Salvatore et al. (2007) | USA | 1989-1995 | FEP | Acute intoxication, withdrawal syndrome, delirium, previous psychiatric hospitalization, unless for detoxification, presence of intellectual disability, non-psychiatric causes of psychiatric symptoms, index syndromal illness present >6 months or previous syndromal episode, prior total treatment with an antipsychotic > 4 weeks or mood-stabilizer for >3 months | Admitted patients | Fair | Y | University health center | 377 | 224/59 | 30.8 | 100 |
| Russell (1994) | USA | NR | Up to age 13, with onset of DSM-III schizophrenia before age 11 | Intellectual disability or known neurologic or  medical disorder affecting the central nervous system | Chart review screening records at one institution, followed by contacting eligible patients and their families | Poor | Y | University health center | 35 | 24/69 | 9.45 | 85.71 |
| Ropcke & Eggers (2005) | Germany | 1979-1988 | SZ spectrum | Affective psychosis | Re-contacting patients after admission | Fair | Y | University health center | 39 | 20/51 | 16.9 | 35.9 |
| Shah et al. (2017) ^c^ | Canada | 2003-2013 | FEP, age 14-35, no antipsychotic medication > 30 days | Intellectual disability, psychotic illness solely related to substance intoxication or withdrawal, or medical or neurological mental disorder. Patients with a concurrent substance use disorder were not excluded. | Patients recruited from a FEP clinical service | Good | Y | University health center | 351 | 248/71 | 23.35 | 67.81 |
| Renwick et al. (2015) | Ireland | 2005-2011 | FEP, age 16-65 | Intellectual disability, psychosis due to another medical condition | Inpatients and outpatients in an early intervention in psychosis program | Good | Y | FEP Program | 375 | 219/58 | 32.9 | 80.53 |
| Perkins et al. (2000) | USA | NR | Clinically stable patients with psychosis | NR | NR | Poor | Y | University health center | 35 | 22/63 | 29 | 85.71 |
| Rabe-Jabllonska et al. (2000) | Poland | 1984-1996 | FEP, age 15-19 | NR | Admitted patients | Good | N | University health center | 150 | 72/48 | 16.7 | 78 |
| Naqvi et al. (2014) | Pakistan | NR | SZ | Comorbid substance abuse or non-psychiatric mental  disorder | Convenience sampling of patients with SZ | Poor | Y | University health center | 93 | 55/59 | NR | 86.02 |
| Mustonen et al. (2018) | Finland | 2001-2002 | All persons consenting from a birth cohort | Psychosis diagnosis before follow-up year (< age 15-16) | Birth cohort: all live born children from the two northernmost provinces in Finland | Fair | Y | Birth cohort | 154 | NR | NR | 52.6 |
| Morgan et al. (2006) | UK | NR | FEP, age 16-65 | Psychotic symptoms precipitated by a non-psychiatric cause, previous treatment for psychosis, transient psychotic symptoms resulting from acute intoxication | All FEP patients reporting to study clinics in 2 catchment areas | Fair | Y | Urban clinics | 470 | 286/61 | 30 | 79.36 |
| Moller & Husby (2000) | Norway | 1994-1996 | FEP, SZ and SZ-phreniform age 18-30, no more than 2 years since first treatment | Medical illness or intellectual disability | Admitted patients | Poor | N | Hospital | 19 | 11/58 | 22.4 | 100 |
| Maki et al. (2014) | Finland | 2001-2008 | National registry: all persons who provided consent | Diagnosis of any psychiatric disorder before 2003, developmental disorders, only substance use/only non-psychiatric disorders during follow-up period | Birth cohort for all of Finland, July 1 1985- June 30 1986 | Fair | Y | National registry | 23 | 13/57 | NR | 60.87 |
| Meng et al. (2009) | Switzerland, Germany, Austria | 1999-2002 | FEP, early onset | Psychopathological syndromes related to neurological or systemic disease, intellectual disability | Admitted patients | Good | Y | Multicenter, university and community sites | 87 | 52/60 | 16.7 | 96.55 |
| Kohn et al. (2004) | Germany | NR | SZ | Intellectual disability | Patients presenting to a number of clinics in a large catchment area | Fair | Y | Urban clinics | 82 | 60/73 | 29.8 | 89.02 |
| Kim et al. (2009) | South Korea | NR | FEP, age 17-45, SZ spectrum or BAD | History of head trauma, comorbid CNS disorder, moderate to severe intellectual disability, transient psychosis after acute intoxication | Admitted patients | Poor | Y | Hospital | 20 | 11/55 | 27.1 | 90 |
| Kanahara et al. (2013) | Japan | 1996-2001 | Psychosis, with no or ineffective previous treatment | Alcohol-or illicit drug-related psychosis, psychosis due to non-psychiatric causes, or psychosis due to dementia | Patients previously admitted and available at 10-year follow-up | Poor | N | Hospital serving a large catchment area | 156 | 77/49 | Not available for the whole sample. Age at onset for "severe cases at admission" 34.2 y (11.9); for "non severe cases at admission" 33.3 y, (12.3) | 73.72 |
| Jackson et al. (1995) | Australia | 1986-1992 | FEP, age </= 45 | Intellectual disability, non-psychiatric causes of symptoms | Admitted patients | Fair | Y | Hospital | 313 | 196/63 | 25.5 | 60%, 44%, 25% |
| Jackson et al. (1996) | Australia | NR | FEP, age 18-45 | Intellectual disability, non-psychiatric causes of symptoms | Patients recruited from a FEP clinical service | Fair | Y | University-affiliated FEP program | 50 | 32/64 | 26.3 | 50 |
| Iida et al. (1995) | Japan | 1984-1993 | SZ, diagnosis age < 15 | NR | Patients presenting to one department of psychiatry | Fair | N | University health center | 39 | 26/67 | 14.62 | 79.49 |
| Huber et al. (1975) | Germany | 1945-1959 | SZ | NR | Admitted patients | Poor | N | Patients interviewed at home or in the hospital | 502 | NR | NR | 46.81 |
| Gourzis et al. (2002) | Greece | 1992-1997 | SZ | NR | All patients admitted to the only inpatient service in a catchment area | Fair | Y | University health center, serving a large catchment area | 100 | 64/64 | 25.6 | 100 |
| Gottlieb (1941) | USA | 1929-1933 | First admission patients with hebephrenic SZ, with a high school education and who were natively urban | No "physical defects" | Admitted patients | Poor | N | Hospital | 100 | NR | NR | 100 |
| Creel (1988) | USA | NR | SZ or SZ-phreniform service members | NR | Random sample from eligible patients in the US Armed Forces | Fair | N | US Armed Forces | 40 | 34/85 | 22.5 | 75 |
| Costello (2012) | USA | 2001-2010 | SZ with at least one related hospital admission or four related outpatient appointments | Schizophrenia cases that remained in active service for more than two years aft er meeting the surveillance case defi nition were assumed to have been misdiagnosed and removed from analysis. Individuals who were considered incident cases of schizophrenia in 2001 were excluded from analyses if they received any schizophrenia diagnoses during the year 2000. | Review of all relevant US Armed Forces records | Fair | N | US Armed Forces | 3000 | 2578/86 | age at dx: 17-24, 60%; 25-29, 24%; 30-34, 10%; 35+ 7% | 71 |
| Coryell & Zimmerman (1988) | USA | NR | Psychosis (delusions, hallucinations, formal thought disorder) | Current mania, medical or drug history which might invalidate the dexamethasone suppression or thyrotropin releasing hormone test | Admitted patients | Poor | Y | University health center, serving a large catchment area | 21 | 9/43 | 33.4 | 66.67 |
| Conus et al. (2007) | Australia | 1998-2000 | FEP | Non-psychotic diagnosis, transferred away from study clinical service | Chart review at a FEP service | Fair | N | University-affiliated FEP program | 597 | 435/73 | 22 | 80.57 |
| Chen et al. (2019) | UK | 2005-2016 | FEP, age 16-45, no antipsychotic medication 1 year prior to FEP diagnosis, registered for 5 years at a participating practice | Parkinson’s disease or dementia in the 5 years prior to FEP, record of psychosis in remission 5 years prior to analysis | National registry of 10 million patients registered with over 670 primary care practices | Fair | N | Primary care practices | 3045 | 1914/63 | median (IQR): 30 (23, 39) | 51.17 |
| Chen et al. (2005) | Hong Kong (China) | 1997-2000 | FEP | Previous psychotic episode, known neurological condition, history of special school attendance (proxy for moderate to severe learning disability). | All patients presenting to public facilities in the catchment area | Fair | Y | Urban public healthcare facilities | 131 | 58/44 | 31.5 | 72.52 |
| Day et al. (1987) | Denmark; India; Colombia; USA; Nigeria; Japan; Czechoslovakia | NR | First psychosis contact, SZ-spectrum diagnosis, onset of florid psychotic symptoms within 7 days, onset of illness occured within 6 months of screening data for study, onset can be dated reliably to within one week's time | NR | NR | Fair | N | Multicenter, urban and rural | 386 | 198/51 | percentage given for each center, see paper if needed. majority under 30; tables 6.1 and 6.2 | 33.42 |
| Bensi et al. (2011) | Italy, UK | 2006-2007 | SZ-spectrum (non-affective), delusional disorder, psychosis NOS, with relapse leading to readmission | NR | Admitted patients | Fair | N | Hospital | 253 | 174/69 | 48.8 | 65.22 |
| Dominguez-Martinez et al. (2017) | Spain | NR | FEP, age 14-40 | NR | Patients recruited from a FEP clinical service | Fair | Y | FEP Program | 40 | 25/63 | 26 | 85 |
| Bechdolf et al. (1998) | Germany | NR | SZ, either first episode or relapse from remission; remitting from symptoms at time of evaluation | NR | Admitted patients | Fair | Y | Psychiatric clinic | 33 | 18/54 | 20 | 96.97 |
| Addington, et al. (2002) | Canada | NR | SZ-spectrum, < 3 months adequate treatment once admitted to the study service, completed 1 year followup | NR | Patients recruited from a FEP clinical service | Fair | Y | University-affiliated FEP program | 86 | 57/66 | 24 | 84.88 |
| Barajas et al. (2019) | Spain | NR | 2 or more psychotic symptoms, age 12-45, initial contact with mental health services within the previous 6 months, < 1 year since onset of psychiatric symptoms | Intellectual disability, head injury, dementia or any non-psychiatric causes of psychosis, low verbal IQ (IQ < 85) | Recruited from adult, child and adolescent community and hospital mental health services in a metropolitan area and outskirts | Poor | Y | Hospitals and community psychiatric services | 79 | 44/56 | 20.22 | 100 |
| Varsamis & Adamson (1971) | Canada | NR | FEP-SZ | NR | Admitted patients | Poor | N | Hospital, urban | 44 | 27/61 | median 31.5 | 75 |
| Hafner et al. (1995 – 2000) ^d^ | Germany | 1987-1989 | First admission for psychosis, diagnosed with SZ broadly, age 12-59 | First admissions age 0-11, non-psychiatric cause of psychosis, severe intellectual disability | Admitted patients, representing 84% of first admissions in the catchment area | Fair | Y | Multisite, urban and rural | 232 | 108/47 | 30.3 | 73.28 |
| Comptom et al. (2006 – 2011) ^e^ | USA | 2004-2008 | Psychosis (non-affective), recent onset or previously untreated, age 18-40, MMSE >/= 23 | Intellectual disability, significant medical condition affecting participation, prior antipsychotic treatment of >3 months, hospitalization for psychosis >3 months prior to index hospitalization | Admitted patients in two public-sector hospitals and an urban county psychiatric crisis center | Fair | Y | Urban, university-affiliated, public-sector hospital and urban county psychiatric crisis center | 109 | 83/76 | 23 | 69.72 |
| Eggers & Bunk (2009) ^f^ | Germany | 1925-1961 | Childhood onset schizophrenia, onset age 7-14 | NR | Admitted patients | Fair | Y | University health center | 57 | NR | NR | 54.39 |
| Guloksuz et al. (2020) | Netherlands | 2007-2016 | Random sample | NR | Random sample | Good | Y | General population sample | 26 | NR | NR | 38.46 |
| Ferrara et al. (2021 | USA | 2014-2019 | Subjects for this analysis were drawn from consecutive admissions to a community health center-based FES (Specialized Treatment Early in Psychosis, STEP) over 5 years (February 1st, 2014, to January 31st, 2019). STEP’s broad eligibility criteria includes between the ages 16–35 who were within the frst 3 years of psychosis onset, and also met Structured Clinical Interview for DSM-IV TR diagnosis for any non-organic schizophrenia-spectrum or schizoafective psychosis, including schizophreniform disorders, brief psychotic episode, and psychosis NOS. Services were also restricted to residents in a ten town catchments contiguous with the clinic in New Haven, Connecticut. | The clinic excluded referrals with an established diagnosis of afective psychosis (Bipolar Disorder and Major Depressive Disorder with psychotic features), and psychosis secondary to substance use or a medical illness. | Subjects for this analysis were drawn from consecutive admissions to a community health center-based FES (Specialized Treatment Early in Psychosis, STEP) over 5 years (February 1st, 2014, to January 31st, 2019). For most of this period, STEP hosted an early detection campaign that sought to recruit a representative sample across the catchment area [21]. | Good | Y | Community, but university affiliated FEP clinic | 168 | 118/70 | 22.4 (3.8) | 88.1 |

*Note.* ^a^ Studies include: Schultze-Lutter et al. (2010), Schultze-Lutter et al. (2015).

^b^ Studies include Schothorst et al. (2006), Emck et al. (2001).

^c^ Studies include: Shah et al. (2017), Pierre (2010), Iyer et al. (2008), Cupo et al. (2021).

^d^ Studies include: Hafner (1996), Maurer et al. (1998), Hafner et al. (1995), Hafner et al. (1999), Hafner (2000), Hafner (1998), Hafner et al. (1999).

^e^ Studies include Compton et al. (2006), Compton et al. (2008), Compton et al. (2009), Compton et al. (2010), Compton et al. (2011).

^f^ Studies include Eggers & Bunk (2009), Eggers & Bunk (1997), Eggers et al. (2003), Eggers (1973), Eggers et al. (2000).
