## Supplementary material for "On the Proportion of Patients Who Experience a Prodrome Prior to Psychosis Onset - A Systematic Review and Meta-analysis": Table 2: Prodrome Definitions

Prodrome definitions: studies and sample sizes

| Type of prodrome definition | Author(s) | Sample size | Total |
| --- | --- | --- | --- |
| Non-specific | Yildizhan et al. (2015) | 43 | 2132 |
|  | Tan & Ang (2001) | 30 |  |
|  | Sullivan (1932) | 100 |  |
|  | Russell (1994) | 35 |  |
|  | Ropcke & Eggers (2005) | 39 |  |
|  | Rabe-Jabllonska et al. (2000) | 150 |  |
|  | Naqvi et al. (2014) | 93 |  |
|  | Morgan et al. (2006) | 470 |  |
|  | Moller & Husby (2000) | 19 |  |
|  | Kohn et al. (2004) | 82 |  |
|  | Kanahara et al. (2013) | 156 |  |
|  | Creel (1988) | 40 |  |
|  | Coryell & Zimmerman (1988) | 21 |  |
|  | Conus et al. (2007) | 597 |  |
|  | Chen et al. (2005) | 131 |  |
|  | Dominguez-Martinez et al. (2017) | 40 |  |
|  | Addington, et al. (2002) | 86 |  |
| Specified Broad | Sandeep (2012) | 51 | 10,823 |
|  | Yung & McGorry (1996) | 21 |  |
|  | Skokou et al. (2012) | 87 |  |
|  | Shioiri et al. (2007) | 219 |  |
|  | Schultze-Lutter et al. (2015) | 126 |  |
|  | Schothorst et al. (2006) | 129 |  |
|  | Stepniak et al. (2014) | 1011 |  |
|  | Salvatore et al. (2007) | 377 |  |
|  | Renwick et al. (2015) | 375 |  |
|  | Perkins et al. (2000) | 35 |  |
|  | Mustonen et al. (2018) | 154 |  |
|  | Maki et al. (2014) | 23 |  |
|  | Meng et al. (2009) | 87 |  |
|  | Kim et al. (2009) | 20 |  |
|  | Jackson et al. (1995) | 313 |  |
|  | Jackson et al. (1996) | 50 |  |
|  | Iida et al. (1995) | 39 |  |
|  | Huber et al. (1975) | 502 |  |
|  | Gourzis et al. (2002) | 100 |  |
|  | Gottlieb (1941) | 100 |  |
|  | Costello (2012) | 3000 |  |
|  | Chen et al. (2019) | 3045 |  |
|  | Day et al. (1987) | 386 |  |
|  | Bensi et al. (2011) | 253 |  |
|  | Bechdolf et al. (1998) | 33 |  |
|  | Barajas et al. (2019) | 79 |  |
|  | Varsamis & Adamson (1971) | 44 |  |
|  | Hafner et al. (1995 – 2000) | 232 |  |
|  | Compton et al. (2006 – 2011) | 109 |  |
|  | Eggers & Bunk (2009) | 57 |  |
| Attenuated (or subthreshold) Psychotic Symptoms | Woodberry et al. (2014) | 40 | 585 |
|  | Shah et al. (2017) | 351 |  |
|  | Guloksuz et al. (2020) | 26 |  |
|  | Ferrara et al (2021) | 168 |  |
