## Supplementary material for "On the Proportion of Patients Who Experience a Prodrome Prior to Psychosis Onset - A Systematic Review and Meta-analysis": Table 3: Subgroup Analyses

Table 3: Meta-analysis Results for Subgroups

| Subgroup | Number of studies included | Sample size (total patients) | Prodrome Prevalence Estimate (Random Effects) | Prodrome Prevalence Estimate 95% CI | I^2^ | I^2^ 95% CI | 95% Predictive Interval |
| --- | --- | --- | --- | --- | --- | --- | --- |
| Only “good” or “fair” quality studies | 37 | 12401 | 76.4% | 70.1%-82% | 98.2% | 97.9%-98.4% | 39.7%-92.8% |
| Only studies conducted at FES/FEP services | 9 | 2001 | 78.1% | 70.2%-85% | 92.9% | 88.7%-95.5% | 44.1%-93.4% |
| Only self-report from patients | 3 | 228 | 54.8% | 48.3%-61.1% | 0% | 0%-92.8% | Not calculated;between study variance estimated at 0 |
| Only studies which conducted interviews | 41 | 6535 | 80.3% | 73.7%-86.2% | 97.5% | 97.1%-97.8% | 38.4%-95.9% |
| Studies which used the DSM-III | 5 | 585 | 81.1% | 46.5%-99.4% | 98.5% | 97.8%-99% | 4.8%-99.8% |
| Studies which used the IRAOS | 8 | 868 | 81.4% | 70.1%-90.5% | 93.5% | 89.5%-96% | 37.4%-96.4% |
| Only studies of large populations or involving catchment areas | 18 | 9069 | 77.7% | 70.5%-84.1% | 97.9% | 97.4%-98.3% | 43.2%-92.4% |
| Only studies which used a prodrome scale | 32 | 3933 | 80.6% | 73.3%-87.0% | 96.6% | 95.9%-97.2% | 41.8%-95.5% |
| Only studies which did not use a prodrome scale | 19 | 9841 | 73.1% | 64.6%-80.8% | 98.5% | 98.2%-98.8% | 31.5%-92.4% |
| Only studies of inpatients | 24 | 3238 | 78.6% | 66.9%-88.3% | 98.2% | 97.8%-98.5% | 32.8%-95.4% |
| Only studies using solely chart review | 6 | 6971 | 74.2% | 59.4%-86.6% | 99.2% | 98.9%-99.4% | 22.1%-95.6% |
| Only studies including mixed diagnosis samples | 23 | 6880 | 77.7% | 67.9%-86.1% | 98.6% | 98.3%-98.8% | 36%-94.2% |
| Only studies including schizophrenia spectrum samples | 28 | 6894 | 78.7% | 72.1%-84.7% | 97% | 96.3%-97.5% | 40.7%-94.5% |
| Only studies using the APS only definition | 4 | 585 | 75.4% | 56%-90.6% | 94.6% | 89.3%-97.3% | 2.7%-99.7& |
| Only studies using the Non-specific definition | 17 | 2132 | 78.6% | 70.8%-85.4% | 93.4% | 90.8%-95.2% | 39.7%-94.7% |
| Only studies using the Specified Broad Definition | 30 | 11057 | 78.4% | 70.9-85.1% | 98.6% | 98.4%-98.8% | 39.9%-93.2% |
